# Effectiveness of SARS-CoV-2 Vaccines against Omicron Infection and Severe Events: A Systematic Review and Meta-Analysis of Test-Negative Design Studies

**DOI:** 10.1101/2023.02.16.23286041

**Authors:** Shangchen Song, Zachary J. Madewell, Mingjin Liu, Ira M. Longini, Yang Yang

**Author notes:** Correspondence to: Dr. Yang Yang, Department of Statistics, University of Georgia, 310 Herty Drive, Athens, GA 30602, USA. Co-first authors (Shangchen Song, Zachary J. Madewell). Co-senior authors (Ira M. Longini, Yang Yang).

## Abstract

**Background:** Evaluating vaccine effectiveness (VE) of a full vaccine series and booster doses against COVID-19 is important for health decision-making.

**Methods:** We systematically searched papers that evaluated VE of SARS-CoV-2 vaccines on PubMed, Web of Science, Cochrane Library, Google Scholar, Embase, Scopus, and preprint servers (bioRxiv and medRxiv) published from November 26th, 2021 to June 27th, 2022 (for full doses and first booster), and to January 8th, 2023 (for the second booster). The pooled VE against Omicron-associated symptomatic or any infection as well as severe events are estimated in a meta-analysis framework.

**Results:** From 2,552 citations identified, a total of 42 were included. The vaccination of the first booster provided stronger protection against Omicron than the full doses alone, shown by the VE estimates of 53.1% (95% CI: 48.0−57.8) vs. 28.6% (95% CI: 18.5−37.4) against infection and 82.5% (95% CI: 77.8−86.2) vs. 57.3% (95% CI: 48.5−64.7) against severe disease. The second booster offered strong protection among adults within 60 days of vaccination against infection (VE=53.1%, 95% CI: 48.0−57.8) and severe disease (VE=87.3% (95% CI: 75.5-93.4), comparable to the first booster with corresponding VE estimates of 59.9% against infection and 84.8% against severe disease. The VEs of the booster doses against severe disease among adults sustained beyond 60 days, 77.6% (95% CI: 69.4−83.6) for the first and 85.9% (95% CI: 80.3−89.9) for the second booster. The VE against infection was less sustainable regardless of dose type. Pure mRNA vaccines provided comparable protection to partial mRNA vaccines, but both provided higher protection than non-mRNA vaccination.

**Conclusion:** One or two booster doses of current SARS-CoV-2 vaccines provide considerable protection against Omicron infection and substantial and sustainable protection against Omicron-induced severe clinical outcomes.

**Funding:** US CDC U01 CK000670

## Introduction

The Omicron variant (B.1.1.529) was first detected in early November 2021 in South Africa and was designated the fifth variant of concern by the World Health Organization.^1^ In contrast to the original wild-type variant, Omicron accumulated over 50 mutations in the whole genome, including 26-32 in the spike protein. This altered protein receptor-binding efficiency and immunogenicity, increasing infectivity, ability to evade neutralizing antibodies, and risk of reinfection.^2^ Additional mutations led to multiple Omicron subvariants with increased transmissibility including BA.2, BA.2.12.1, BA.4, BA.4.6, BA.5, BF.7, BQ.1, BQ.1.1, and XBB.1.5, the latter three of which accounted for most infections in the United States as of February, 2023.^3^ The effective reproduction number (R_t_) and basic reproduction number (R_0_) were estimated to be 3.8 and 2.5 times higher for Omicron than for Delta.^4^ Compared with the wild-type and Delta variants, Omicron replicates less efficiently in the lung parenchymal tissues and more efficiently in the bronchial tissues, which may contribute to increased transmissibility but decreased disease severity.^5–7^

There is a rapidly growing body of literature of real-world vaccine effectiveness (VE) against Omicron. Studies reported that individuals vaccinated with two mRNA doses were less susceptible to Omicron infection, though the level of protection conferred was lower than that of earlier variants, and protection waned over time.^8,9^ The emergence of new variants coupled with waning vaccine-induced immunity prompted recommendations for booster doses and second booster doses based on the original vaccine formula, which were shown to confer greater protection against Omicron than two mRNA doses.^10,11^ Omicron-specific bivalent mRNA booster doses were recently authorized for use in the U.S. by the Food and Drug Administration, and early data demonstrated stronger neutralizing antibody responses against Omicron than the original monovalent mRNA vaccines.^12^ The BNT162b2 bivalent BA.4/5 COVID-19 vaccine was recently shown to elicit greater neutralizing antibody titers against newer Omicron sublineages (BA.4.6, BA.2.75.2, BQ.1.1 and XBB.1) in adults older than 55 than a fourth dose of the original monovalent BNT162b2.^13^ Uptake of the bivalent boosters, however, is low with only 15% of the U.S. adult population vaccinated as of February 2023.^14^ Therefore, it is important to quantify the effectiveness of the original vaccines against Omicron.

Two early meta-analyses evaluated VE of a primary vaccine series or single booster dose and demonstrated greater protection for the third dose against symptomatic infection and severe events compared to a two-dose regimen.^15,16^ However, they focused on hybrid immunity (immunity developed from SARS-CoV-2 infection and vaccination)^15^ and relative vaccine effectiveness of the third dose compared to two doses^16^ rather than non-vaccination. Nor did they evaluate VE for a second booster, long-term (>60 days) VE for the first booster, or adult- and child-specific VEs. Herein, we aggregate estimates in the literature to evaluate VE for the initial full doses, first booster dose, and second booster dose against Omicron-related infection and severe events for pure mRNA, partial (mixed) mRNA, and non-mRNA vaccines. We focus our review on test-negative design studies, an increasingly popular epidemiological study design for evaluating VE on infectious pathogens including influenza, rotavirus, pneumococcus, and others.^17^ In this design, the same clinical definition is used to enroll cases and controls and laboratory testing distinguishes “test positive” cases from “test negative” controls, thereby reducing bias from differential healthcare-seeking behavior between cases and controls.^18^

## Methods

### Data sources, eligibility criteria, search strategies, and data extraction

This analysis followed the Preferred Reporting Items for Systematic Reviews and Meta-analyses (PRISMA) reporting guidelines. A systematic literature search was conducted of PubMed, Web of Science, Cochrane Library, Google Scholar, Embase, Scopus, and preprint servers (bioRxiv and medRxiv) for papers published from November 26th, 2021, when Omicron was classified as a World Health Organization Variant of Concern,^1^ to June 27th, 2022 (for full doses and booster), and to January 8th, 2023 (for the second booster).

The selection of studies followed Participant (P), Intervention (I), Comparator (C), Outcome (O), and Study Type (S), PICOS criteria^19^ (supplementary materials p 2). Published studies were eligible for inclusion if they were original analyses with the test-negative design and reported VE or corresponding odds ratios (OR) of full doses, booster, or second booster against Omicron infection or severe events. We excluded studies that focused on special populations (e.g., patients with kidney disease); did not include circulation period of Omicron variant; combined VE estimates for Omicron with other viral variants such as Delta; reported relative VE between different vaccines, vaccination doses, or variants among vaccinated individuals; did not evaluate VE (e.g., instead, evaluated neutralizing antibodies); or evaluated outcomes other than infection or severe events. All available ages were included. We did not contact authors for additional data.

We applied Boolean combinations of the following keywords to identify relevant publications: “SARS-CoV-2”, “COVID-19”, “2019nCoV”, “vaccine”, “booster”, “second booster”, “effectiveness”, “efficacy”, “test-negative case-control”, “test-negative design”, “Omicron”, “infection”, “hospitalization”; the detailed search procedures were presented in the supplementary materials. Publication language was not restricted, and reference lists of selected papers were also screened for additional studies.

After removing duplicated results, we first screened studies by titles and abstracts to identify potentially eligible articles. Two pairs of researchers then independently evaluated full texts and selected those meeting the inclusion criteria. Any disagreements were discussed until a consensus was reached. Preprints were checked and updated with their most recent published version if available as of January 10^th^, 2023. Zotero was used for literature management. Finally, two pairs of researchers independently extracted the following from the included studies: author names, publication year, study region, study design, dose, vaccine type, test time in reference to vaccination time, adjusted VE point estimate and 95% confidence intervals, and adjustment confounders; if available, the number of vaccinated and unvaccinated individuals in the cases and controls were also recorded.

### Evaluation of study quality and risk of bias

Study quality and risk of bias were independently assessed by two researchers using the Newcastle-Ottawa Scale (NOS). Studies could earn up to 9 points composed of participant selection (4 points), study comparability (1 point), and outcome of interest (4 points). A score >7 was considered as high quality, 5–6 as medium, and <5 as low, and studies classified as low were excluded from the meta-analysis. Publication bias was also evaluated by Egger’s test, Begg and Mazumdar rank correlation, and funnel plots when at least ten studies were available, with significance set at *p* < 0.1. If we detected publication bias, we used the Duval and Tweedie trim-and-fill method^20^ for adjustment, which consists of imputing missing effect sizes to achieve symmetry.

### Statistical analysis

We categorized full doses and booster VE into short-term, long-term, and overall to evaluate potential waning of VE over time. There is no uniform definition for short-term vs. long-term VE, but most studies adopted cut-off points of 60-120 days from last vaccination to lab-testing. We used these cut-off points to guide the classification of VE estimates into short-term vs. long-term (supplementary materials p 2). If a study reported VEs for finer time intervals than we needed, we used an inverse variance-weighted (IVW) averaging approach to combine them.

For each time interval, we further categorized VE by the type of vaccine: pure mRNA vaccines, partial mRNA vaccines, and non-mRNA vaccines. Pure mRNA vaccines comprise of homogenous or heterogeneous BNT162b2 and mRNA-1273, or a population-level mixture of the two if a study does not discriminate between them. Partial mRNA vaccines include either a multi-dose course containing at least one mRNA vaccine dose, or the study indiscriminately reported VEs of a population-level mixture of vaccines including at least one mRNA vaccine. Non-mRNA vaccines refers to the regimens that do not involve mRNA vaccines at all (e.g., Ad26.COV2.S, ChAdOx1).

We evaluated VE against Omicron infection and severe events. Analyses of VE against infection or symptomatic infection combined studies that reported either VE against symptomatic infection or VE against any infection (symptomatic or asymptomatic). Severe events included hospitalizations, noncritical hospitalizations, deaths, emergency department (ED) or urgent care (UC) encounters, ED admissions, intensive care unit (ICU) admissions, and invasive ventilation.

We evaluated VE for the overall vaccine-eligible population as well as for age groups defined as adults (≥18 years) and children/adolescents (5-17 years). If VE was not reported but odds ratios (OR) were provided, we calculated VE as (1 − OR) ×100%. The pooled VE and 95% confidence intervals were calculated via a random effects meta-analysis with restricted maximum likelihood estimation. *I*^*2*^ was used to evaluate between-study heterogeneity with thresholds of 25%, 50%, and 75% indicating low, moderate, and high heterogeneity, respectively. The metafor package in the R statistical software (version 4.0.5) was used for estimation and visualization in this meta-analysis.^21^

## Results

For full doses and booster doses, we obtained 1,139 articles from all searched databases (82 from PubMed, 23 from Web of Science, 89 from Embase, 721 from Scopus, 3 from Cochrane Library, 115 from medRxiv, 6 from bioRxiv, and 100 from Google Scholar). After removing duplicates, 952 articles remained, of which 136 were retained for full review following inspection of the title, abstract, and keywords. After full text review of these 136 articles, 33^9,22–53^ with 271 VE estimates were formally included in this meta-analysis (supplementary materials p 18). For the second booster, we obtained 1,413 articles from all databases (56 from PubMed, 22 from Web of Science, 55 from Embase, 1,015 from Scopus, nine from Cochrane Library, 149 from medRxiv, seven from bioRxiv, and 100 from Google Scholar). After removing duplicates, 1,236 articles remained, of which 116 were considered relevant after inspection of the title, abstract, and keywords. These 116 relevant articles were then reviewed in full text for eligibility, and 11 articles^23,37,54–62^ with 46 VE estimates were finally included in this meta-analysis (supplementary materials p 19). More summary details and detected publication bias of the included studies are given in the supplementary materials (p 2).

The VE estimates for the initial full doses against Omicron symptomatic infection or any infection were summarized in Figure 1. Pooling all vaccine types and time intervals, the overall VE was estimated to be 28.6% (95% CI: 18.5-37.4%, 25 studies) for all ages and 24.4% (95% CI: 16.2-31.8%, 15 studies) for adults. Overall VE of the pure mRNA vaccines was estimated to be 30.6% (95% CI: 17.1-41.8%, 18 studies) for all ages, 25.4% (95% CI: 11.5-37.1%, 8 studies) for adults, and 54.2% (95% CI: 35.2-67.7%, 5 studies) for children and adolescents. Overall VE estimates for partial mRNA vaccines and non-mRNA vaccines were only available for adults, 28.1% (95% CI: 19.8-35.6%, 5 studies) and 1.5% (95% CI: 0.4-2.7%, 2 studies) respectively. This is also why we do not have a separate overall VE estimate for children and adolescents pooling all vaccine types.

**Figure 1.**
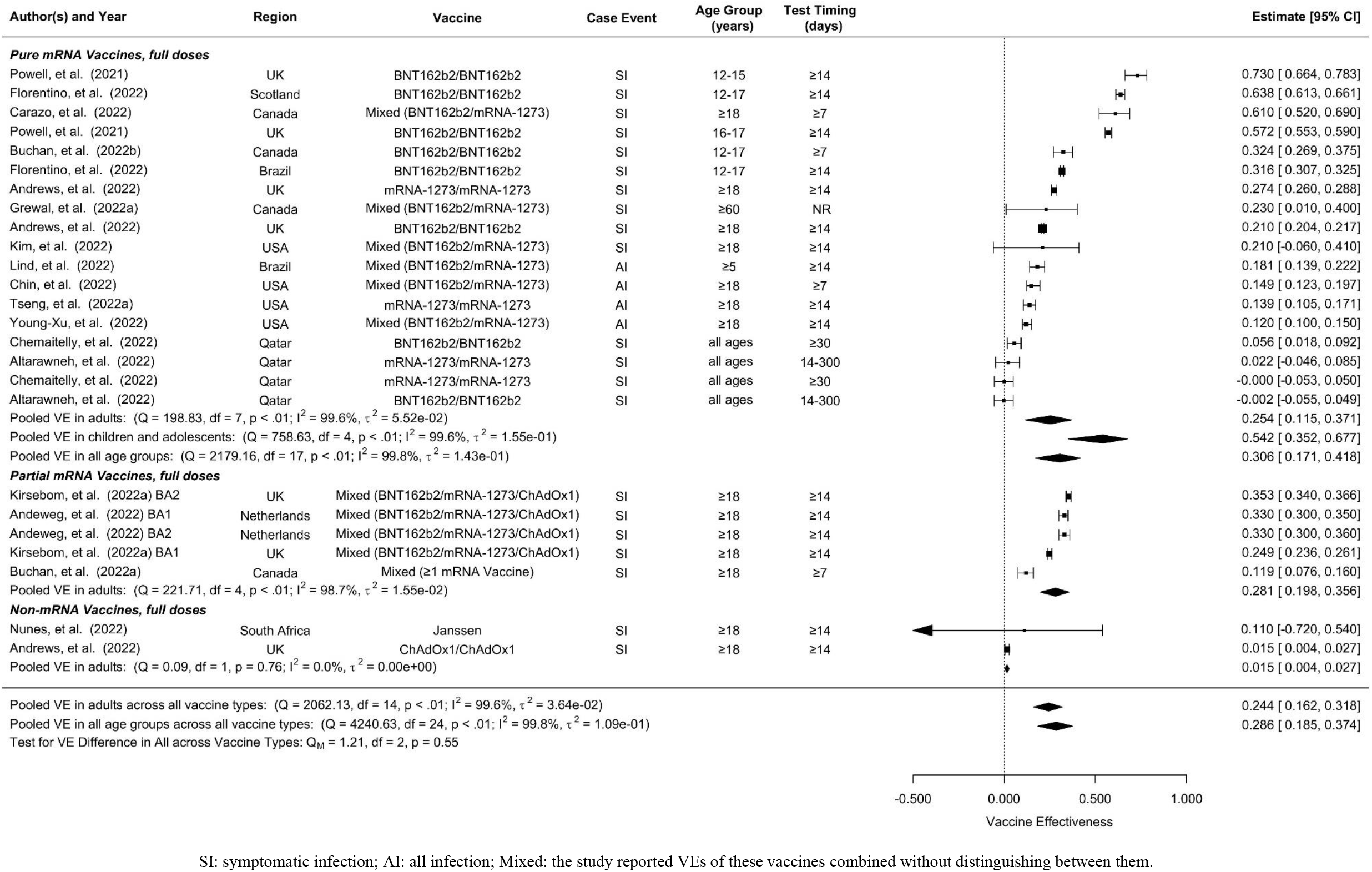
Overall vaccine effectiveness of full dose against infection or symptomatic infection. Pooled VE is estimated from all 25 studies combined as well as for each vaccine type. Statistics Cochran’s Q, I^2^ and τ^2^ measure the heterogeneity between studies. End points of the studies are either symptomatic infection (SI) or any infection (AI). Mixed vaccine type indicates the study reported VEs of these vaccines combined without distinguishing between them.

Short-term full-dose VE estimates pooling all vaccine types were 40.7% (95% CI: 34.3-46.5%, 19 studies) for all ages and 37.5% (95% CI: 31.4-43.1%, 10 studies) for adults (supplementary materials p 3). Short-term VE of pure mRNA vaccines was estimated to be 43.5% (95% CI: 35.4-50.6%, 13 studies) for all ages, 41.3% (95% CI: 40.2-42.4%, 4 studies) for adults, and 45.3% (95% CI: 28.7-58.1%, 6 studies) for children and adolescents. Short-term VE estimate of partial mRNA vaccines was 34.7% (95% CI: 25.4-42.9%, 6 studies) for adults, slightly lower than that of the pure mRNA vaccines.

Long-term full-dose VE estimates against symptomatic or any infection were in general much lower than their short-term counterparts. Pooling all vaccine types, long-term full-dose VE was estimated to be 17.6% (95% CI: 13.2-21.8%, 22 studies) for all ages and 16.6% (95% CI: 10.5-22.3%, 15 studies) for adults (supplementary materials p 4). Long-term full-dose VE of pure mRNA vaccines was estimated to be 16.4% (95% CI: 13.6-19.1%, 11 studies) for all ages, 13.1% (95% CI: 11.7-14.6%, 4 studies) for adults, and 22.3% (95% CI: 13.6-30.1%, 4 studies) for children and adolescents. Long-term full-dose VE among adults was estimated to be 22.6% (95% CI: 10.8-32.7%, 5 studies) for partial mRNA vaccines and 13.2% (95% CI: 2.6-22.6%, 6 studies) for non-mRNA vaccines.

Compared to unvaccinated controls, the overall VE of the first booster dose against Omicron symptomatic infection or any infection was 53.1% (95% CI: 48.0-57.8%, 31 studies) for all ages and 53.4% (95% CI: 47.7-58.6%, 27 studies) for adults (Figure 2). No studies included in this analysis reported VE of booster doses for children. When stratified by vaccine type, the overall first-booster VE estimates were 58.0% (95% CI: 51.4-63.6%, 11 studies) for all ages and 61.4% (95% CI: 54.1-67.5%, 7 studies) in adults for pure mRNA vaccination, 56.4% (95% CI: 52.7-59.8%, 15 studies) for adults for partial mRNA vaccines, and 25.2% (95% CI: 2.2-42.8%, 5 studies) for adults for non-mRNA vaccines,

**Figure 2.**
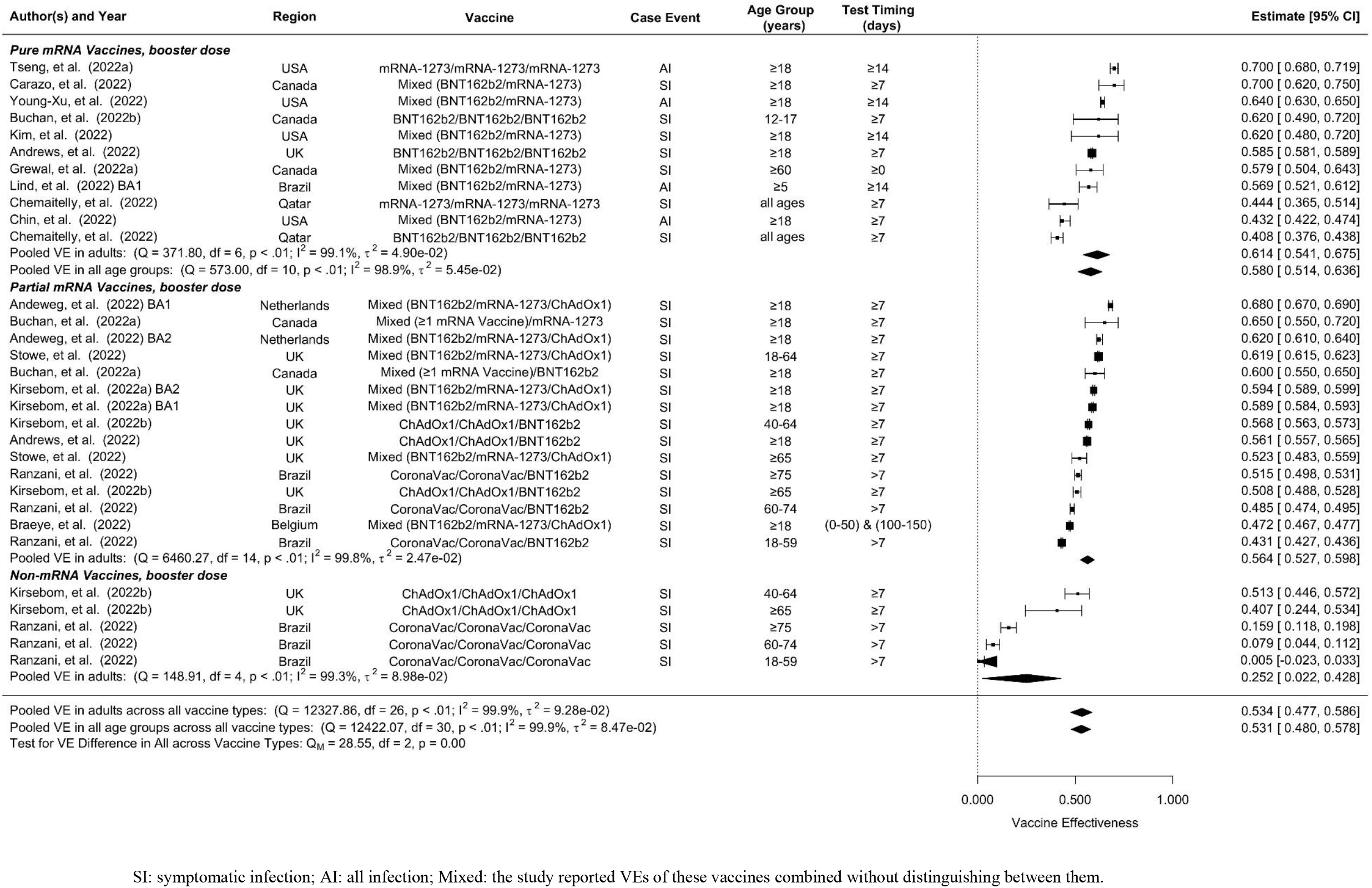
Overall vaccine effectiveness of the first booster dose against infection or symptomatic infection. Pooled VE is estimated from all 31 studies combined as well as for each vaccine type. Statistics Cochran’s Q, I^2^ and τ^2^ measure the heterogeneity between studies. End points of the studies are either symptomatic infection (SI) or any infection (AI). Mixed vaccine type indicates the study reported VEs of these vaccines combined without distinguishing between them.

In comparison to its overall VE, the short-term VE estimates of the first booster dose were slightly higher, 59.4% (95% CI: 55.1-63.3%, 33 studies) for all ages and 59.9% (95% CI: 55.1-64.1%, 28 studies) for adults (supplementary materials p 5). When stratified by vaccine type, the short-term first-booster VE estimates were 63.7% (95% CI: 59.2-67.7%, 15 studies) for all ages and 67.3 % (95% CI: 64.5-69.9%, 10 studies) for adults for pure mRNA vaccination, 62.3% (95% CI: 59.2-65.1%, 12 studies) for adults for partial mRNA vaccines, and 37.2% (95% CI: 19.5-51.0%, 6 studies) for adults for non-mRNA vaccines.

Long-term VE estimates of the first booster dose were moderately lower than their overall counterparts, 34.9% (95% CI: 27.6-41.5%, 22 studies) for all ages and 31.5% (95% CI: 22.7-39.4%, 20 studies) for adults (supplementary materials p 6). Long-term first-booster VE estimates stratified by vaccine type were 46.6% (95% CI: 36.8-54.8%, 7 studies) for all ages and 50.9% (95% CI: 45.0-56.2%, 5 studies) for adults for pure mRNA vaccination, 34.6% (95% CI: 28.6-40.2%, 11 studies) for adults for partial mRNA vaccines, and 4.6% (95% CI: -9.5-16.9%, 4 studies) for adults for non-mRNA vaccines.

Due to lack of data, we were only able to estimate short-term and long-term VE but not overall VE of the second booster (Figure 3). Furthermore, we were unable to distinguish between vaccine types for the second booster, but the majority of these studies were based on four doses of mRNA vaccines. The short-term second-booster VE against symptomatic infection or any infection for Omicron was 59.6% (95% CI: 52.0-66.1%, 17 studies) in adults, similar to the overall and the short-term first-booster VE estimates in adults. The long-term second-booster VE was 32.7% (95% CI: 15.4-46.4%, 10 studies) in adults, comparable to that of the first booster.

**Figure 3.**
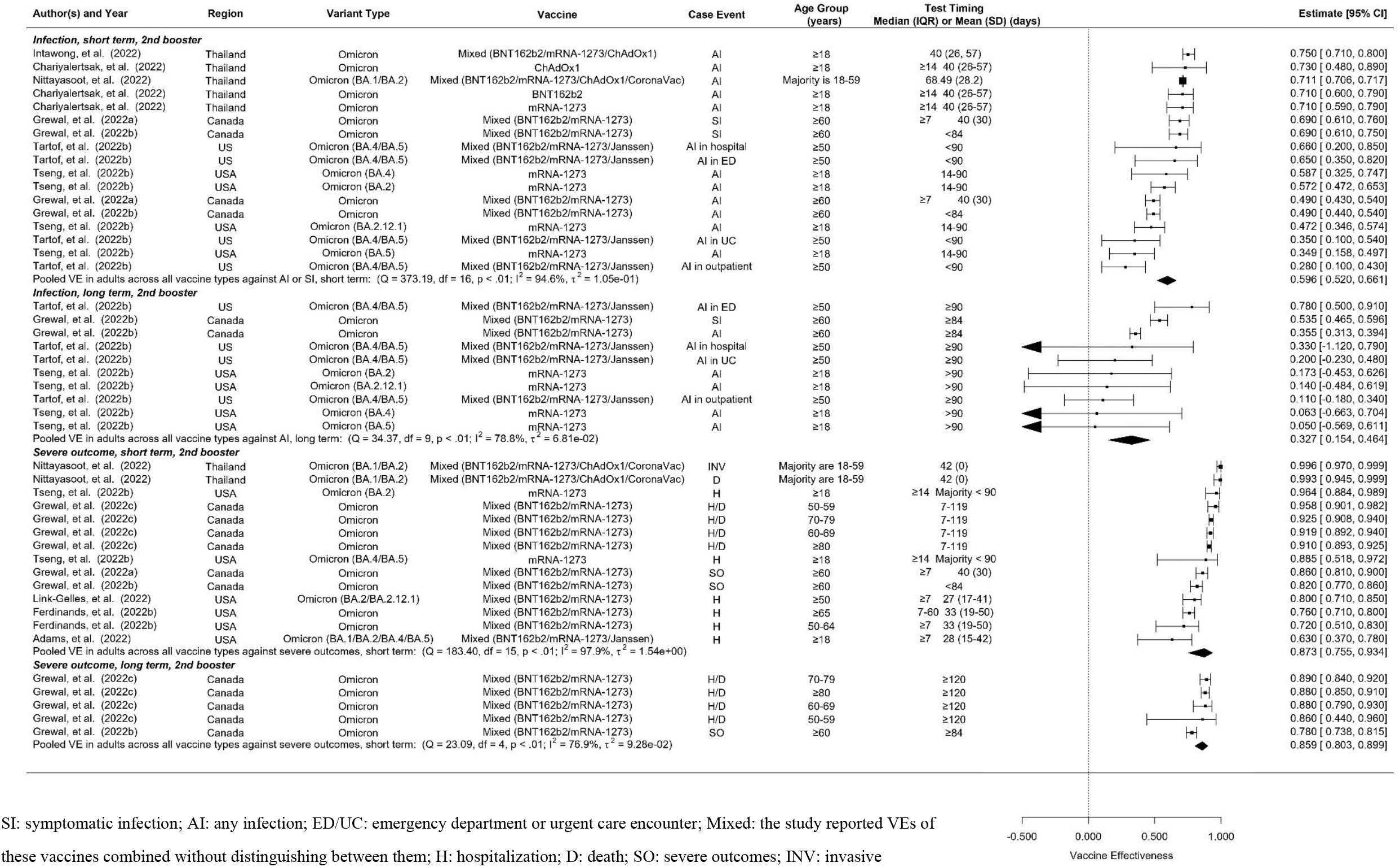
Overall vaccine effectiveness of the second booster dose against infection or symptomatic infection and against severe events. Pooled VE estimates are stratified by short-term (<60 days) vs. long-term (≥60 days). Statistics Cochran’s Q, I^2^ and τ^2^ measure the heterogeneity between studies. For infection, possible end points of the studies are symptomatic infection (SI) or any infection (AI). For severe events, possible end points are hospitalization (H), death (D), severe outcomes (SO) or invasive procedures (INV). Mixed vaccine type indicates the study reported VEs of these vaccines combined without distinguishing between them.

Overall VE of the full doses against Omicron-associated severe events was estimated to be 57.3% (95% CI: 48.5%-64.7%, 24 studies) for all ages and 57.9% (95% CI: 51.5%-63.4%, 16 studies) for adults (Figure 4). Overall VE estimates of pure mRNA vaccines were 60.9% (95% CI: 50.7-68.9%, 18 studies) for all ages, 60.1% (95% CI: 53.1-66.0%, 10 studies) for adults, and 59.9% (95% CI: 24.7-78.6%, 6 studies) for children and adolescents. Overall VE of partial mRNA vaccines for adults was slightly lower than that of pure mRNA vaccines, 54.5% (95% CI: 41.1-64.8%, 6 studies). We did not find studies estimating the overall VE of non-mRNA vaccines against Omicron-related severe events.

**Figure 4.**
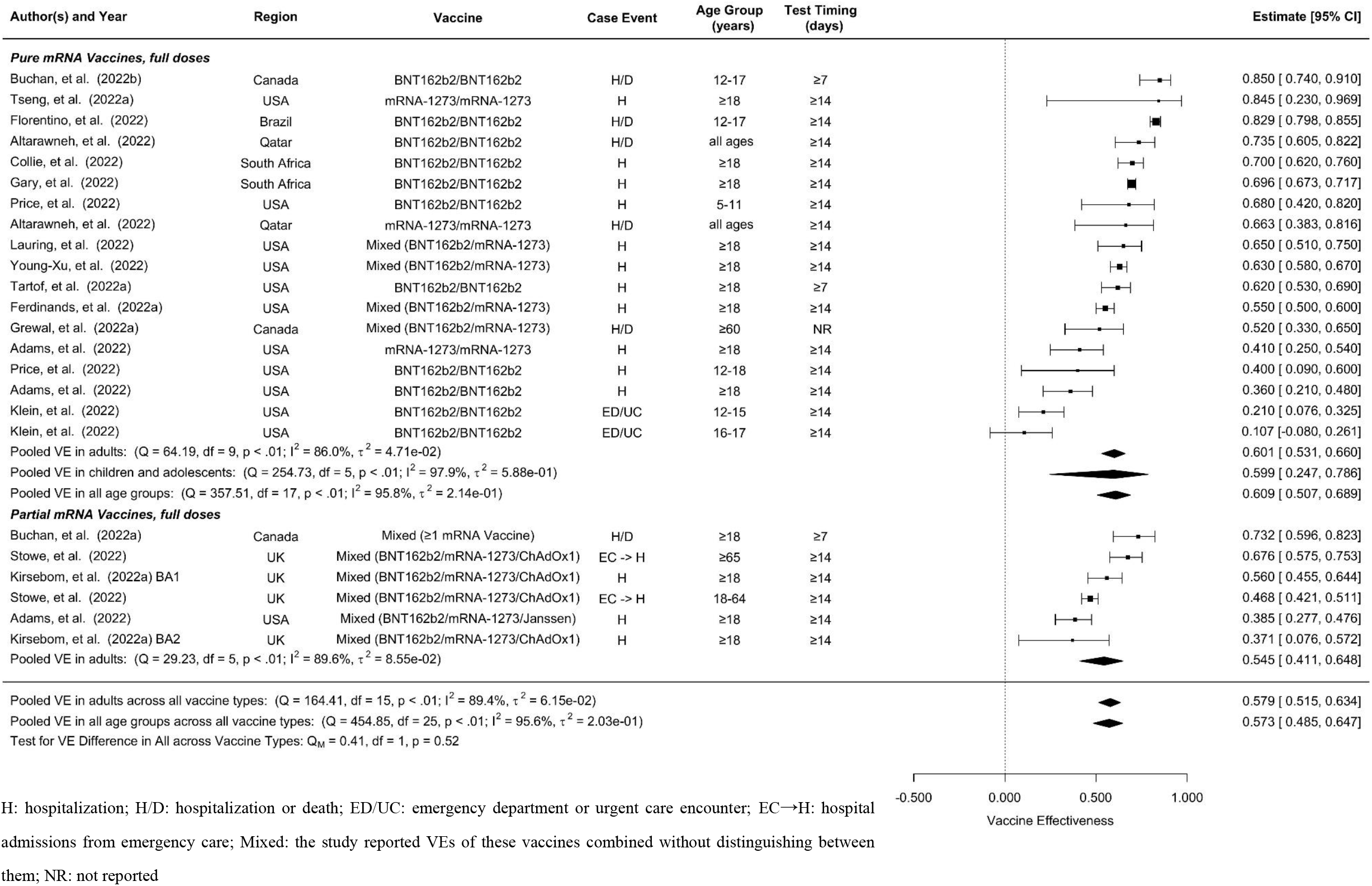
Overall vaccine effectiveness of full dose against severe events. Pooled VE is estimated from all 24 studies combined as well as for each vaccine type. Statistics Cochran’s Q, I^2^ and τ^2^ measure the heterogeneity between studies. Possible end points of the studies are hospitalization (H), hospitalization or death (H/D), emergency department or urgent care encounter (ED/UC), or hospital admissions from emergency care (EC→H). Mixed vaccine type indicates the study reported VEs of these vaccines combined without distinguishing between them.

Short-term VE of the full doses against Omicron-associated severe events was estimated to be 66.9% (95% CI: 58.3-73.8%, 16 studies) for all ages and 69.9% (95% CI: 62.8-75.6%, 10 studies) for adults (supplementary materials p 7). Stratified by vaccine type, the short-term VE estimates were 64.0% (95% CI: 50.2-74.0%, 9 studies) for all ages, 70.5% (95% CI: 64.9-75.2%, 3 studies) for adults, 60.7% (95% CI: 36.6%-75.6%, 6 studies) for children and adolescents for pure mRNA vaccines and 70.7% (95% CI: 59.2%-78.9%, 7 studies) for adults for partial mRNA vaccines.

Long-term VE estimates of the full doses against Omicron-associated severe events were comparable to the overall VE estimates, 58.3% (95% CI: 45.5-68.1%, 18 studies) for all ages and 59.0% (95% CI: 49.0-67.1%, 13 studies) for adults (supplementary materials p 8). Stratified by vaccine type, the long-term VE estimates were 62.4% (95% CI: 38.9-76.8%, 9 studies) for all ages, 67.7% (95% CI: 56.3-76.1%, 4 studies) for adults, and 56.4% (95% CI: -3.6-81.7%, 5 studies) for children and adolescents for pure mRNA vaccines, 50.7% (95% CI: 29.9-65.2%, 6 studies) for adults for partial mRNA vaccines, and 60.1% (95% CI: 39.7-73.6%, 3 studies) for adults for non-mRNA vaccines.

First booster doses generally showed higher VEs against Omicron-associated severe disease than full doses. The pooled overall VE of the first booster dose was estimated to be 82.5% (95% CI: 77.8%-86.2%, 28 studies) for all ages and 82.0% (95% CI: 77.0%-86.0%, 25 studies) for adults (Figure 5). Pure mRNA vaccines and partial mRNA vaccines showed similar overall VEs against severe events, 83.6% (95% CI: 77.0-88.2%, 11 studies) for all ages, 82.5% (95% CI: 74.7-88.0%, 8 studies) for adults for the former, and 84.6% (95% CI: 77.6%-89.5%, 12 studies) for adults for the latter. Overall VE was moderately lower for non-mRNA vaccines, 71.4% (95% CI: 52.1-82.9%, 5 studies) for adults.

**Figure 5.**
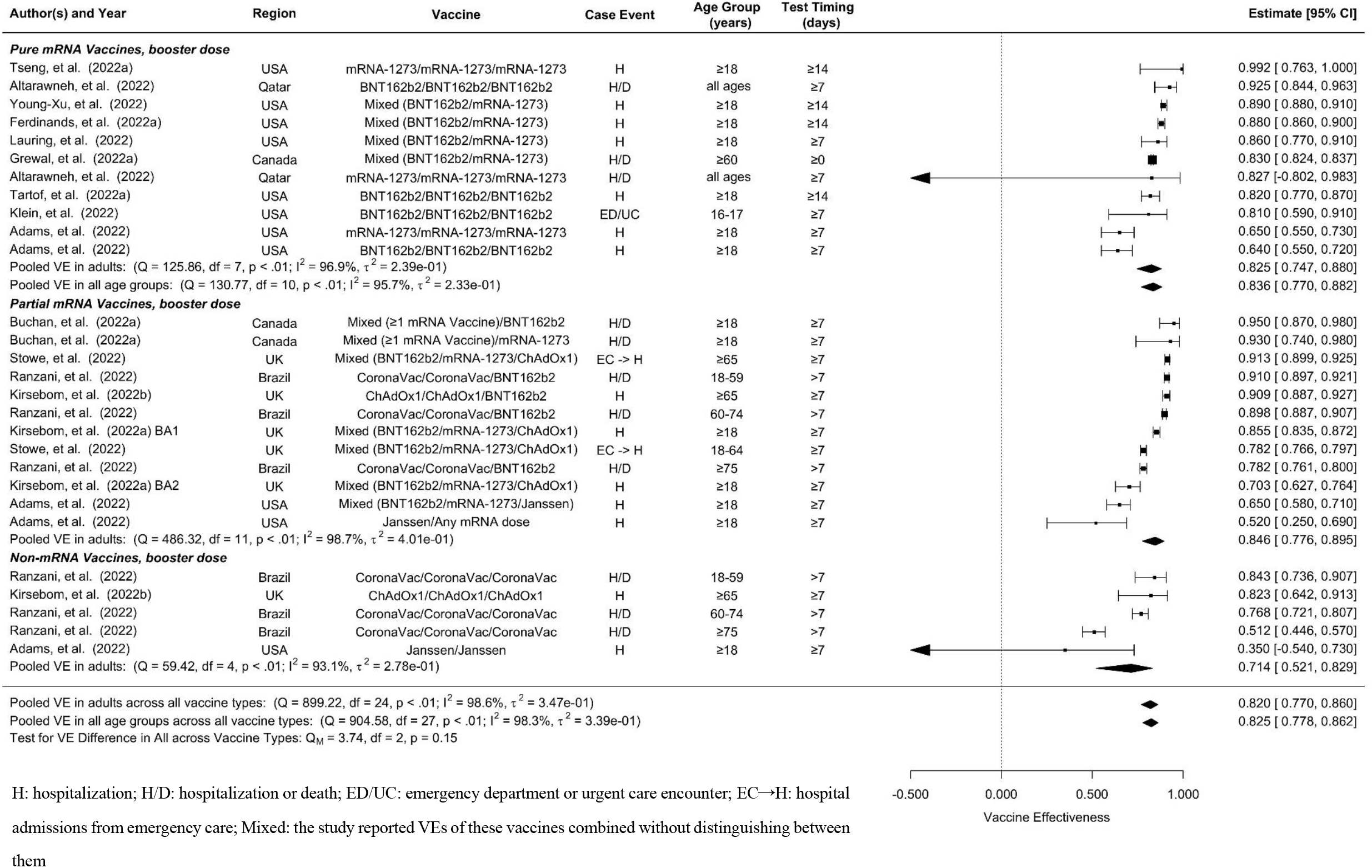
Overall vaccine effectiveness of the first booster dose against severe events. Pooled VE is estimated from all 28 studies combined as well as for each vaccine type. Statistics Cochran’s Q, I^2^ and τ^2^ measure the heterogeneity between studies. Possible end points of the studies are hospitalization (H), hospitalization or death (H/D), emergency department or urgent care encounter (ED/UC), or hospital admissions from emergency care (EC→H). Mixed vaccine type indicates the study reported VEs of these vaccines combined without distinguishing between them.

Short-term and long-term VEs of the booster dose against Omicron-associated severe events were only available for adults (supplementary materials p 9). We estimated short-term VE to be 84.8% (95% CI: 80.4-88.1%, 17 studies) and long-term VE to be 77.6% (95% CI: 69.4-83.6%, 16 studies) for all vaccine types combined. Short-term vs. long-term booster VE estimates were 85.3% (95% CI: 79.8%-89.3%, 6 studies) vs. 80.1% (95% CI: 64.6-88.8%, 5 studies) for pure mRNA vaccines, 88.1% (95% CI: 83.4-91.4%, 7 studies) vs. 78.0% (95% CI: 64.3-86.4%, 8 studies) for partial mRNA vaccines, and 73.0% (95% CI: 53.7-84.3%, 4 studies) vs. 70.5% (95% CI: 47.3-83.5%, 3 studies) for non-mRNA vaccines.

Pooled short-term and long-term VE estimates for the second booster against Omicron-associated severe events among adults were 87.3% (95% CI: 75.5-93.4%, 14 studies), and 85.9% (95% CI: 80.3-89.9%, 5 studies) respectively (Figure 3), both of which are comparable to those of the first booster, though the long-term VE of the second booster appears to decay at a slower rate.

## Discussion

In this systematic review and meta-analysis of 42 studies, we found that one or two booster doses in addition to the initial full COVID-19 vaccine series provided substantial protection against Omicron infection with VE ≥ 50% and severe events with VE ≥ 80%, compared to no vaccination. In general, pure and partial mRNA vaccines provided comparable protection levels against infection or severe disease, and both were more effective than non-mRNA vaccines, though the difference was less dramatic in terms of protection against severe disease. The VEs of the full doses and the booster doses against severe disease only wane slightly after three months, but the VEs against infection wane more quickly.

Both the first and second booster doses provided considerably higher VE against infection and severe events compared to completion of the initial full series only. Studies have reported higher anti-receptor binding domain specific memory B cells and anti-spike antibodies after booster doses compared to full series only.^23,63^ Similarly, T cell immunity against Omicron is provided by booster doses though at a reduced level compared to ancestral variants.^64^ While the initial full doses provided inadequate protection against infection (Figure 1), they did render practically meaningful (≥50%) VE against severe disease (Figure 4).

Pure and partial mRNA vaccines offered comparable protection levels against infection, 25.4% vs. 28.1% for the full doses and 61.4% vs. 56.4% for the first booster among adults, and both were much more effective than the non-mRNA vaccines (1.5% for the full doses and 25.2% for the first booster). Studies included in this analysis reported lower binding activities between anti-spike and anti-receptor among Ad26.COV2 recipients compared to mRNA recipients.^23^ Similar trends were observed against severe events, though the gap between mRNA and non-mRNA vaccines was much narrower. In particular, full-dose non-mRNA vaccines provided a similar level of sustained protection against severe disease (VE=60%) compared to full-dose mRNA vaccines (supplementary materials p 8), suggesting that the initial full doses of non-mRNA vaccines should be encouraged among unvaccinated individuals in regions where mRNA vaccine supply is insufficient.

The VEs of the initial full doses and the first booster dose against Omicron infection waned substantially over time, from 40.7% within three months of boosting to 17.6% for full doses and 59.4% to 34.9% for the first booster. The VEs against Omicron-associated severe disease waned at a slower pace, from 66.9% to 58.3% for the full doses and from 84.8% to 77.6% (in adults) for the first booster dose. Our findings are consistent with other studies reporting waning immunity of COVID-19 vaccines for earlier variants^19, 59^ as well as for Omicron regardless of age, immunocompromised status, and vaccine product.^56^ One study reported that VE against symptomatic infection waned more rapidly among older adults,^65^ which was also reflected in this meta-analysis, e.g., the full-dose VE of pure mRNA vaccines against infection declined from 45.3% to 22.3% among children and from 41.3% to 13.1% among adults (supplementary materials pp 3-4). These age differences in decay rates were not observed for the VEs against severe disease (supplementary materials pp 7-8).

The second booster of pure or partial mRNA vaccines protected adults from Omicron infection with a VE of 59.6% which is slightly lower than the short-term VE of the first booster for pure mRNA (67.3%) or partial mRNA vaccines (62.3%) among adults. A similar gap was seen for the long-term VE among adults as well, 32.7% for the second booster vs. 50.9% for pure mRNA and 34.6% for partial mRNA first boosters. This seemingly unexpected gap (not statistically significant) may result from the fact that the dominant Omicron subvariants were mostly BA.1 and BA.2 for the first booster studies but BA.4 and BA.5 were taking over for the second booster studies. BA.4 and BA.5 are known to be associated with high immune escape and transmissibility compared to BA.1 and BA.2, e.g., the effective reproductive number was estimated to be 5.11 and 5.22 for BA.4 and BA.5 compared to 3.22 and 5.04 for BA.1 and BA.2.^66^

In terms of protection against severe disease among adults, we observed comparable VE estimates between the second booster and the first booster doses for both short term (87.3% for the second booster vs. 85.3% and 88.1% for pure and partial mRNA first boosters) and long term (85.9% for second booster vs. 80.1% and 78.0 for pure and partial mRNA first boosters). The second booster appears to wane to a lesser extent over time. However, a caveat is that nearly all data used to estimate the long-term VE of the second booster against severe disease came from the same study among elderly residents of long-term care facilities in Ontario, Canada.^61^ In addition, this long-term VE is against BA.1 and BA.2, the dominant subvariants during the study period of 31 Dec 2021 to 27 April 2022, according to the Ontario Ministry of Health.

Our study had several limitations. First, in several test-negative studies, we included, the same control group for multiple vaccine groups, which introduces dependence among the VE estimates. However, such dependence was not accounted for in our analysis due to lack of covariance estimates. Second, there was significant heterogeneity in VE estimates, which may be attributable to differences between studies in terms of a whole host of characteristics, including study design, follow-up duration, definitions of VE, time since vaccination, dosing intervals, confounders adjusted for, and others.

Our findings demonstrate that completion of a full COVID-19 vaccine series plus one or two booster doses provides considerable VE against Omicron infection and strong VE against severe events compared to non-vaccination. Although VEs generally wane after 2-3 months, the second booster clearly generates more sustainable protection. To facilitate comparison and synthesis of VE estimates across studies, we recommend the following improvements to future vaccine studies: (1) longer follow-up to better understand long-term VE; (2) stratification of VE by age group and vaccine type whenever possible; and (3) when multiple VE estimates are reported, providing covariance or correlation among the estimates via, e.g., resampling the data.

## Data Availability

All data produced in the present study are available upon reasonable request to the authors

## Contributor

YY and IL conceived the study. SS, ZM and ML collected the data, SS, ZM, ML and YY reviewed the data. SS analyzed data under the supervision of YY, ZM and IL. SS, ZM and YY drafted the manuscript. All authors contributed to interpretation of results and critical revision.

## Acknowledgements

YY, IL and SS were supported by the US CDC grant (U01 CK000670).

