## Supplemental 1 for "Effectiveness of SARS-CoV-2 Vaccines against Omicron Infection and Severe Events: A Systematic Review and Meta-Analysis of Test-Negative Design Studies"

### Supplementary Materials

#### Supplementary methods

*Determination of short-term and long-term VE*

We categorized full doses and booster VE into short-term, long-term, and overall to evaluate potential waning of VE over time. There is no uniform definition for short-term vs. long-term VE. In addition, studies reported VE by post-vaccination stage using different time intervals, but most studies adopted cut-off points of 60-120 days. Considering the lower and upper bounds of the post vaccination test dates, we used the following guidelines. For initial full doses, a lower bound ≤30 days and an upper bound ≤180 days constitute short term, and a lower bound ≥90 (except one study used ≥70 days) days and an upper bound that is either ≥200 days or unspecified are considered long term. For booster doses, a lower bound ≤30 days and an upper bound ≤120 days are considered short term, and a lower bound ≥60 days and an upper bound >120 days or unspecified are considered as long-term. To simplify description, we occasionally use "<90 days" and "≥90 days" to represent short-term vs. long-term VEs for the full doses, and use "<60 days" and "≥60 days" to represent short-term vs. long-term VEs for the booster doses.

#### Supplementary Results

*Summary of studies included*

For full doses and booster doses, 33 papers [9,22–53] were formally included in this meta-analysis (sFigure 13, appendix p 18). There were 14 studies conducted in the U.S., five in the U.K., four in Canada, three in South Africa, two in Qatar, two in Brazil, and one in each of Belgium, Netherlands and Scotland, respectively. A study could report multiple VEs for different vaccination types and outcomes. In total, there were 271 VE estimates including 124 for full doses and 147 for the first booster doses; 133 for pure mRNA vaccines, 100 for partial mRNA vaccines, and 38 the non-mRNA vaccines; 138 for symptomatic infection, 14 for any infection, and 119 for severe events. For the second booster, 11 articles[23,37,54–62] were finally included in this meta-analysis (sFigure 14, appendix p 19), including five studies conducted in the U.S., three in Canada, and three in Thailand. In total, there were 46 VE estimates including 32 for pure mRNA vaccines, 13 for partial mRNA vaccines, and one for non-mRNA vaccines; three for symptomatic infection, 24 for any infections, and 19 for severe events.

*Publication bias*

Publication bias was detected in the pooled estimates of overall VE of the full doses against severe events (Egger's test $p$ = 0·073, Begg's test $p$ = 0·208), long-term VE of the full doses against severe events (Egger's test $p$ = 0·027, Begg's test $p$ = 0·369), short-term VE of the first booster dose against severe events (Egger's test $p$ = 0·098, Begg's test $p$ = 0·49), and short-term VE of the second booster dose against severe events (Egger's test $p$ = 0·001, Begg's test $p$ = 0·747), as shown in supplementary figure 8-11 (appendix pp 11-14). Additionally, publication bias was found in four subgroups defined by age group and vaccine type (appendix, sFigure 12, p 15). Results were corrected for these biases using the trim-and-fill method.

#### Supplementary Tables

**sTable 1 PICOS Criteria**

| | |
|---|---|
| Participant | The general population excluding disease-specific cohorts. |
| Intervention | Full doses, booster or second booster COVID-19 vaccinations |
| Comparator | Unvaccinated participants |
| Outcome | VE=1-adjusted odds ratio estimated in a (conditional) logistic regression |
| Study Type | Test-negative design study |

#### Supplementary Figure

| Author(s) and Year | Region | Vaccine | Case Event | Age Group (years) | Test Timing (days) | Estimate [95% CI] |
|---|---|---|---|---|---|---|
| ***Pure mRNA Vaccines, full doses*** | | | | | | |
| Florentino, et al. (2022) | Scotland | BNT162b2/BNT162b2 | SI | 12-17 | 14-97 | 0.648 [0.623, 0.671] |
| Powell, et al. (2021) | UK | BNT162b2/BNT162b2 | SI | 16-17 | 14-69 | 0.629 [0.610, 0.646] |
| Chemaitelly, et al. (2022) | Qatar | BNT162b2/BNT162b2 | SI | all ages | 30-119 | 0.478 [0.408, 0.539] |
| Kim, et al. (2022) | USA | Mixed (BNT162b2/mRNA-1273) | SI | ≥18 | 14-149 | 0.450 [0.140, 0.660] |
| Tseng, et al. (2022a) | USA | mRNA-1273/mRNA-1273 | AI | ≥18 | 14-90 | 0.440 [0.351, 0.516] |
| Chemaitelly, et al. (2022) | Qatar | mRNA-1273/mRNA-1273 | SI | all ages | 30-119 | 0.432 [0.150, 0.621] |
| Andrews, et al. (2022) | UK | mRNA-1273/mRNA-1273 | SI | ≥18 | 14-104 | 0.423 [0.400, 0.445] |
| Andrews, et al. (2022) | UK | BNT162b2/BNT162b2 | SI | ≥18 | 14-104 | 0.410 [0.400, 0.419] |
| Buchan, et al. (2022b) | Canada | BNT162b2/BNT162b2 | SI | 12-17 | 7-119 | 0.378 [0.293, 0.452] |
| Fleming-Dutra, et al. (2022) | USA | BNT162b2/BNT162b2 | SI | 5-11 | (14-30) & (60-90) | 0.362 [0.326, 0.396] |
| Florentino, et al. (2022) | Brazil | BNT162b2/BNT162b2 | SI | 12-17 | 14-97 | 0.341 [0.332, 0.351] |
| Lind, et al. (2022) | Brazil | Mixed (BNT162b2/mRNA-1273) | AI | ≥5 | 14-149 | 0.285 [0.200, 0.362] |
| Fleming-Dutra, et al. (2022) | USA | BNT162b2/BNT162b2 | SI | 12-15 | (14-30) & (60-90) | 0.215 [0.139, 0.285] |
| Pooled VE in adults: ($Q = 1.72$, $df = 3$, $p = 0.63$; $I^2 = 9.3\%$, $\tau^2 = 6.50e-05$) | | | | | | 0.413 [0.402, 0.424] |
| Pooled VE in children and adolescents: ($Q = 781.29$, $df = 5$, $p < .01$; $I^2 = 99.4\%$, $\tau^2 = 1.09e-01$) | | | | | | 0.453 [0.287, 0.581] |
| Pooled VE in all age groups: ($Q = 826.53$, $df = 12$, $p < .01$; $I^2 = 99.2\%$, $\tau^2 = 5.48e-02$) | | | | | | 0.435 [0.354, 0.506] |
| ***Partial mRNA Vaccines, full doses*** | | | | | | |
| Kirsebom, et al. (2022a) BA2 | UK | Mixed (BNT162b2/mRNA-1273/ChAdOx1) | SI | ≥18 | 14-175 | 0.449 [0.431, 0.466] |
| Andeweg, et al. (2022) BA2 | Netherlands | Mixed (BNT162b2/mRNA-1273/ChAdOx1) | SI | ≥18 | 30-119 | 0.430 [0.351, 0.499] |
| Andeweg, et al. (2022) BA1 | Netherlands | Mixed (BNT162b2/mRNA-1273/ChAdOx1) | SI | ≥18 | 14-119 | 0.412 [0.359, 0.460] |
| Kirsebom, et al. (2022a) BA1 | UK | Mixed (BNT162b2/mRNA-1273/ChAdOx1) | SI | ≥18 | 14-175 | 0.349 [0.334, 0.365] |
| Braeye, et al. (2022) | Belgium | Mixed (BNT162b2/mRNA-1273/ChAdOx1) | SI | ≥18 | 0-100 | 0.208 [0.194, 0.221] |
| Buchan, et al. (2022a) | Canada | Mixed (≥1 mRNA Vaccine) | SI | ≥18 | 7-119 | 0.197 [0.124, 0.264] |
| Pooled VE in adults: ($Q = 488.76$, $df = 5$, $p < .01$; $I^2 = 98.8\%$, $\tau^2 = 2.65e-02$) | | | | | | 0.347 [0.254, 0.429] |
| Pooled VE in adults across all vaccine types: ($Q = 833.10$, $df = 9$, $p < .01$; $I^2 = 98.6\%$, $\tau^2 = 1.97e-02$) | | | | | | 0.375 [0.314, 0.431] |
| Pooled VE in all age groups across all vaccine types: ($Q = 1666.46$, $df = 18$, $p < .01$; $I^2 = 99.3\%$, $\tau^2 = 4.84e-02$) | | | | | | 0.407 [0.343, 0.465] |
| Test for VE Difference in All across Vaccine Types: $Q_M = 1.74$, $df = 1$, $p = 0.19$ | | | | | | |

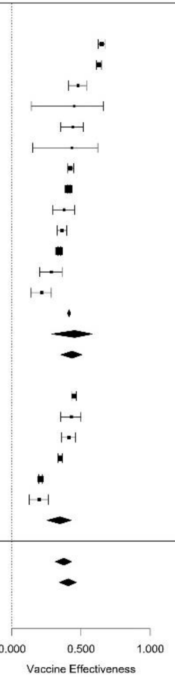

sFigure 1 Short-term vaccine effectiveness of full dose against infection or symptomatic infection

| Author(s) and Year | Region | Vaccine | Case Event | Age Group (years) | Test Timing (days) | Estimate [95% CI] |
|---|---|---|---|---|---|---|
| ***Pure mRNA Vaccines, full doses*** | | | | | | |
| Florentino, et al. (2022) | Scotland | BNT162b2/BNT162b2 | SI | 12-17 | ≥98 | 0.313 [ 0.048, 0.505] |
| Buchan, et al. (2022b) | Canada | BNT162b2/BNT162b2 | SI | 12-17 | ≥120 | 0.290 [ 0.216, 0.357] |
| Powell, et al. (2021) | UK | BNT162b2/BNT162b2 | SI | 16-17 | ≥70 | 0.226 [ 0.145, 0.299] |
| Chemaitelly, et al. (2022) | Qatar | mRNA-1273/mRNA-1273 | SI | all ages | 120-419 | 0.187 [ 0.113, 0.255] |
| Chemaitelly, et al. (2022) | Qatar | BNT162b2/BNT162b2 | SI | all ages | 120-419 | 0.163 [ 0.097, 0.225] |
| Lind, et al. (2022) | Brazil | Mixed (BNT162b2/mRNA-1273) | AI | ≥5 | ≥150 | 0.153 [ 0.104, 0.200] |
| Andrews, et al. (2022) | UK | mRNA-1273/mRNA-1273 | SI | ≥18 | ≥140 | 0.150 [ 0.118, 0.181] |
| Florentino, et al. (2022) | Brazil | BNT162b2/BNT162b2 | SI | 12-17 | ≥98 | 0.139 [ 0.109, 0.169] |
| Tseng, et al. (2022a) | USA | mRNA-1273/mRNA-1273 | AI | ≥18 | 181-270 | 0.138 [ 0.102, 0.173] |
| Andrews, et al. (2022) | UK | BNT162b2/BNT162b2 | SI | ≥18 | ≥105 | 0.126 [ 0.118, 0.134] |
| Kim, et al. (2022) | USA | Mixed (BNT162b2/mRNA-1273) | SI | ≥18 | ≥150 | 0.110 [-0.210, 0.350] |
| Pooled VE in adults: (Q = 2.44, df = 3, p = 0.49; $I^2$ = 20.4%, $\tau^2$ = 8.14e-05) | | | | | | 0.131 [ 0.117, 0.146] |
| Pooled VE in children and adolescents: (Q = 16.98, df = 3, p < .01; $I^2$ = 78.7%, $\tau^2$ = 7.94e-03) | | | | | | 0.223 [ 0.136, 0.301] |
| Pooled VE in all age groups: (Q = 30.71, df = 10, p < .01; $I^2$ = 77.0%, $\tau^2$ = 1.66e-03) | | | | | | 0.164 [ 0.136, 0.191] |
| ***Partial mRNA Vaccines, full doses*** | | | | | | |
| Andeweg, et al. (2022) BA2 | Netherlands | Mixed (BNT162b2/mRNA-1273/ChAdOx1) | SI | ≥18 | 150-209 | 0.337 [ 0.299, 0.373] |
| Andeweg, et al. (2022) BA1 | Netherlands | Mixed (BNT162b2/mRNA-1273/ChAdOx1) | SI | ≥18 | 150-209 | 0.301 [ 0.274, 0.327] |
| Kirsebom, et al. (2022a) BA2 | UK | Mixed (BNT162b2/mRNA-1273/ChAdOx1) | SI | ≥18 | ≥175 | 0.278 [ 0.259, 0.297] |
| Kirsebom, et al. (2022a) BA1 | UK | Mixed (BNT162b2/mRNA-1273/ChAdOx1) | SI | ≥18 | ≥175 | 0.148 [ 0.129, 0.167] |
| Buchan, et al. (2022a) | Canada | Mixed (≥1 mRNA Vaccine) | SI | ≥18 | 180-239 | 0.010 [-0.080, 0.100] |
| Pooled VE in adults: (Q = 183.87, df = 4, p < .01; $I^2$ = 98.7%, $\tau^2$ = 2.51e-02) | | | | | | 0.226 [ 0.108, 0.327] |
| ***Non-mRNA Vaccines, full doses*** | | | | | | |
| Ranzani, et al. (2022) | Brazil | CoronaVac/CoronaVac | SI | ≥75 | ≥180 | 0.281 [ 0.247, 0.313] |
| Ranzani, et al. (2022) | Brazil | CoronaVac/CoronaVac | SI | 60-74 | ≥180 | 0.220 [ 0.198, 0.242] |
| Kirsebom, et al. (2022b) | UK | ChAdOx1/ChAdOx1 | SI | ≥65 | ≥175 | 0.195 [ 0.117, 0.266] |
| Kirsebom, et al. (2022b) | UK | ChAdOx1/ChAdOx1 | SI | 40-64 | ≥175 | 0.080 [ 0.060, 0.099] |
| Ranzani, et al. (2022) | Brazil | CoronaVac/CoronaVac | SI | 18-59 | ≥180 | 0.008 [-0.004, 0.020] |
| Andrews, et al. (2022) | UK | ChAdOx1/ChAdOx1 | SI | ≥18 | ≥175 | -0.027 [-0.042, -0.012] |
| Pooled VE in adults: (Q = 500.14, df = 5, p < .01; $I^2$ = 99.4%, $\tau^2$ = 2.02e-02) | | | | | | 0.132 [ 0.026, 0.226] |
| Pooled VE in adults across all vaccine types: (Q = 1399.28, df = 14, p < .01; $I^2$ = 99.2%, $\tau^2$ = 1.80e-02) | | | | | | 0.166 [ 0.105, 0.223] |
| Pooled VE in all age groups across all vaccine types: (Q = 1448.84, df = 21, p < .01; $I^2$ = 98.6%, $\tau^2$ = 1.38e-02) | | | | | | 0.176 [ 0.132, 0.218] |
| Test for VE Difference in All across Vaccine Types: $Q_M$ = 2.76, df = 2, p = 0.25 | | | | | | |

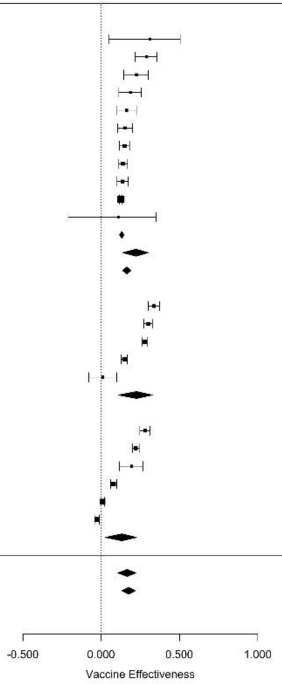

sFigure 2 Long-term vaccine effectiveness of full dose against infection or symptomatic infection

| Author(s) and Year | Region | Vaccine | Case Event | Age Group (years) | Test Timing (days) | Estimate [95% CI] |
|---|---|---|---|---|---|---|
| ***Pure mRNA Vaccines, booster dose*** | | | | | | |
| Andrews, et al. (2022) | UK | BNT162b2/BNT162b2/mRNA-1273 | SI | ≥18 | 7-69 | 0.725 [0.720, 0.731] |
| Accorsi, et al. (2022) | USA | mRNA-1273/mRNA-1273/mRNA-1273 | SI | ≥18 | 14-90 | 0.720 [0.690, 0.740] |
| Tseng, et al. (2022a) | USA | mRNA-1273/mRNA-1273/mRNA-1273 | AI | ≥18 | 14-60 | 0.716 [0.697, 0.734] |
| Fleming-Dutra, et al. (2022) | USA | BNT162b2/BNT162b2/BNT162b2 | SI | 12-15 | 14-45 | 0.711 [0.655, 0.757] |
| Andrews, et al. (2022) | UK | mRNA-1273/mRNA-1273/mRNA-1273 | SI | ≥18 | 7-34 | 0.672 [0.654, 0.689] |
| Accorsi, et al. (2022) | USA | Mixed (BNT162b2/mRNA-1273) | SI | ≥18 | 14-90 | 0.670 [0.650, 0.690] |
| Accorsi, et al. (2022) | USA | BNT162b2/BNT162b2/BNT162b2 | SI | ≥18 | 14-90 | 0.650 [0.620, 0.680] |
| Andrews, et al. (2022) | UK | mRNA-1273/mRNA-1273/BNT162b2 | SI | ≥18 | 7-34 | 0.646 [0.628, 0.663] |
| Andrews, et al. (2022) | UK | BNT162b2/BNT162b2/BNT162b2 | SI | ≥18 | 7-69 | 0.625 [0.621, 0.630] |
| Grewal, et al. (2022a) | Canada | Mixed (BNT162b2/mRNA-1273) | SI | ≥60 | <84 | 0.620 [0.510, 0.710] |
| Altarawneh, et al. (2022) | Qatar | mRNA-1273/mRNA-1273/mRNA-1273 | SI | all ages | 7-60 | 0.613 [0.533, 0.679] |
| Grewal, et al. (2022a) | Canada | BNT162b2/BNT162b2/mRNA-1273 | SI | ≥60 | 20-110 | 0.570 [0.400, 0.690] |
| Altarawneh, et al. (2022) | Qatar | BNT162b2/BNT162b2/BNT162b2 | SI | all ages | 7-60 | 0.540 [0.504, 0.573] |
| Chemaitelly, et al. (2022) | Qatar | mRNA-1273/mRNA-1273/mRNA-1273 | SI | all ages | 7-41 | 0.471 [0.384, 0.546] |
| Chemaitelly, et al. (2022) | Qatar | BNT162b2/BNT162b2/BNT162b2 | SI | all ages | 7-69 | 0.440 [0.405, 0.473] |
| Pooled VE in adults: ($Q = 756.16$, $df = 9$, $p < .01$; $I^2 = 97.4\%$, $\tau^2 = 1.44e\text{-}02$) | | | | | | 0.673 [0.645, 0.699] |
| Pooled VE in all age groups: ($Q = 1083.00$, $df = 14$, $p < .01$; $I^2 = 98.9\%$, $\tau^2 = 4.82e\text{-}02$) | | | | | | 0.637 [0.592, 0.677] |
| ***Partial mRNA Vaccines, booster dose*** | | | | | | |
| Andeweg, et al. (2022) BA1 | Netherlands | Mixed (BNT162b2/mRNA-1273/ChAdOx1) | SI | ≥18 | 7-59 | 0.690 [0.682, 0.698] |
| Kirsebom, et al. (2022a) BA2 | UK | Mixed (BNT162b2/mRNA-1273/ChAdOx1) | SI | ≥18 | 7-69 | 0.683 [0.676, 0.690] |
| Andrews, et al. (2022) | UK | ChAdOx1/ChAdOx1/mRNA-1273 | SI | ≥18 | 7-69 | 0.677 [0.672, 0.682] |
| Kirsebom, et al. (2022a) BA1 | UK | Mixed (BNT162b2/mRNA-1273/ChAdOx1) | SI | ≥18 | 7-69 | 0.660 [0.655, 0.665] |
| Andeweg, et al. (2022) BA2 | Netherlands | Mixed (BNT162b2/mRNA-1273/ChAdOx1) | SI | ≥18 | 7-59 | 0.634 [0.616, 0.650] |
| Ranzani, et al. (2022) | Brazil | CoronaVac/CoronaVac/BNT162b2 | SI | ≥75 | 8-59 | 0.625 [0.602, 0.647] |
| Kirsebom, et al. (2022b) | UK | ChAdOx1/ChAdOx1/BNT162b2 | SI | ≥65 | 7-69 | 0.606 [0.585, 0.627] |
| Kirsebom, et al. (2022b) | UK | ChAdOx1/ChAdOx1/BNT162b2 | SI | 40-64 | 7-69 | 0.602 [0.597, 0.607] |
| Andrews, et al. (2022) | UK | ChAdOx1/ChAdOx1/BNT162b2 | SI | ≥18 | 7-69 | 0.583 [0.579, 0.588] |
| Ranzani, et al. (2022) | Brazil | CoronaVac/CoronaVac/BNT162b2 | SI | 60-74 | 8-59 | 0.579 [0.565, 0.593] |
| Ranzani, et al. (2022) | Brazil | CoronaVac/CoronaVac/BNT162b2 | SI | 18-59 | 8-59 | 0.573 [0.568, 0.579] |
| Braeye, et al. (2022) | Belgium | Mixed (BNT162b2/mRNA-1273/ChAdOx1) | SI | ≥18 | 0-50 | 0.520 [0.520, 0.530] |
| Pooled VE in adults: ($Q = 3423.84$, $df = 11$, $p < .01$; $I^2 = 99.6\%$, $\tau^2 = 1.86e\text{-}02$) | | | | | | 0.623 [0.592, 0.651] |
| ***Non-mRNA Vaccines, booster dose*** | | | | | | |
| Kirsebom, et al. (2022b) | UK | ChAdOx1/ChAdOx1/ChAdOx1 | SI | 40-64 | 7-69 | 0.536 [0.464, 0.598] |
| Andrews, et al. (2022) | UK | ChAdOx1/ChAdOx1/ChAdOx1 | SI | ≥18 | 7-69 | 0.520 [0.446, 0.583] |
| Kirsebom, et al. (2022b) | UK | ChAdOx1/ChAdOx1/ChAdOx1 | SI | ≥65 | 7-69 | 0.489 [0.334, 0.607] |
| Ranzani, et al. (2022) | Brazil | CoronaVac/CoronaVac/CoronaVac | SI | ≥75 | 8-59 | 0.371 [0.267, 0.460] |
| Ranzani, et al. (2022) | Brazil | CoronaVac/CoronaVac/CoronaVac | SI | 60-74 | 8-59 | 0.122 [0.047, 0.190] |
| Ranzani, et al. (2022) | Brazil | CoronaVac/CoronaVac/CoronaVac | SI | 18-59 | 8-59 | 0.081 [0.046, 0.114] |
| Pooled VE in adults: ($Q = 175.64$, $df = 5$, $p < .01$; $I^2 = 97.2\%$, $\tau^2 = 9.08e\text{-}02$) | | | | | | 0.372 [0.195, 0.510] |
| Pooled VE in adults across all vaccine types: ($Q = 7437.23$, $df = 27$, $p < .01$; $I^2 = 99.8\%$, $\tau^2 = 8.76e\text{-}02$) | | | | | | 0.599 [0.551, 0.641] |
| Pooled VE in all age groups across all vaccine types: ($Q = 7614.05$, $df = 32$, $p < .01$; $I^2 = 99.8\%$, $\tau^2 = 8.33e\text{-}02$) | | | | | | 0.594 [0.551, 0.633] |
| Test for VE Difference in All across Vaccine Types: $Q_M = 30.00$, $df = 2$, $p = 0.00$ | | | | | | |

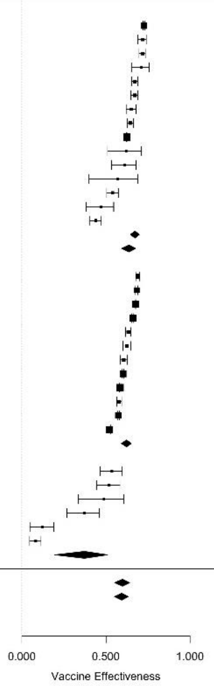

sFigure 3 Short-term vaccine effectiveness of first booster dose against infection or symptomatic infection

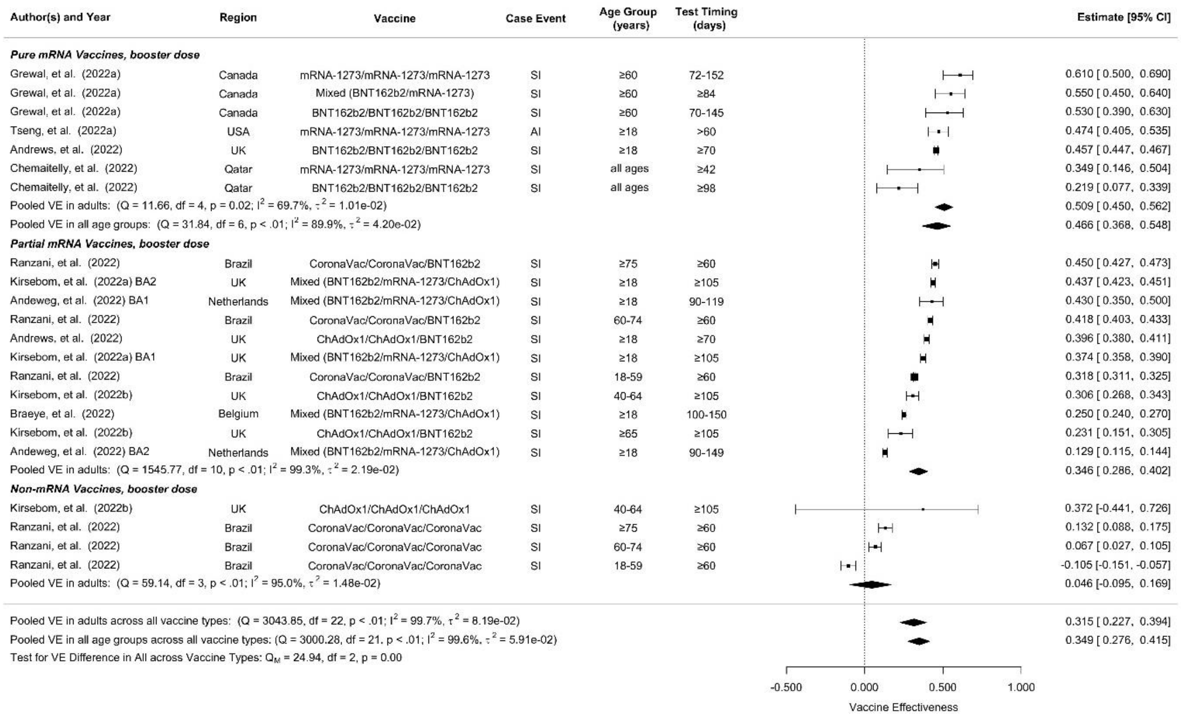

sFigure 4 Long-term vaccine effectiveness of first booster dose against infection or symptomatic infection

| Author(s) and Year | Region | Vaccine | Case Event | Age Group (years) | Test Timing (days) | Estimate [95% CI] |
|---|---|---|---|---|---|---|
| ***Pure mRNA Vaccines, full doses*** | | | | | | |
| Florentino, et al. (2022) | Brazil | BNT162b2/BNT162b2 | H/D | 12-17 | 14-97 | 0.826 [ 0.793, 0.854] |
| Buchan, et al. (2022b) | Canada | BNT162b2/BNT162b2 | H/D | 12-17 | 7-119 | 0.814 [ 0.587, 0.916] |
| Ferdinands, et al. (2022a) | USA | Mixed (BNT162b2/mRNA-1273) | H | ≥18 | <60 | 0.710 [ 0.510, 0.830] |
| Gary, et al. (2022) | South Africa | BNT162b2/BNT162b2 | H | ≥18 | 14-87 | 0.709 [ 0.644, 0.761] |
| Tartof, et al. (2022a) | USA | BNT162b2/BNT162b2 | H | ≥18 | <90 | 0.680 [ 0.480, 0.800] |
| Klein, et al. (2022) | USA | BNT162b2/BNT162b2 | ED/UC | 5-11 | 14-67 | 0.510 [ 0.300, 0.650] |
| Klein, et al. (2022) | USA | BNT162b2/BNT162b2 | ED/UC | 12-15 | 14-149 | 0.450 [ 0.300, 0.570] |
| Price, et al. (2022) | USA | BNT162b2/BNT162b2 | H | 12-18 | 14-160 | 0.430 [-0.010, 0.680] |
| Klein, et al. (2022) | USA | BNT162b2/BNT162b2 | ED/UC | 16-17 | 14-149 | 0.340 [ 0.080, 0.530] |
| Pooled VE in adults: (Q = 0.13, df = 2, p = 0.94; $I^2$ = 0.0%, $\tau^2$ = 0.00e+00) | | | | | | 0.705 [ 0.649, 0.752] |
| Pooled VE in children and adolescents: (Q = 94.01, df = 5, p < .01; $I^2$ = 92.1%, $\tau^2$ = 3.06e-01) | | | | | | 0.607 [ 0.366, 0.756] |
| Pooled VE in all age groups: (Q = 94.81, df = 8, p < .01; $I^2$ = 88.9%, $\tau^2$ = 2.00e-01) | | | | | | 0.640 [ 0.502, 0.740] |
| ***Partial mRNA Vaccines, full doses*** | | | | | | |
| Kirsebom, et al. (2022a) BA2 | UK | Mixed (BNT162b2/mRNA-1273/ChAdOx1) | H | ≥18 | 14-175 | 0.863 [ 0.528, 0.960] |
| Stowe, et al. (2022) | UK | Mixed (BNT162b2/mRNA-1273/ChAdOx1) | EC -> H | ≥65 | 14-174 | 0.808 [ 0.641, 0.897] |
| Natarajan, et al. (2022) | USA | Mixed (Janssen/1mRNADose) | H | ≥18 | 7-120 | 0.780 [ 0.700, 0.840] |
| Kirsebom, et al. (2022a) BA1 | UK | Mixed (BNT162b2/mRNA-1273/ChAdOx1) | H | ≥18 | 14-175 | 0.761 [ 0.593, 0.860] |
| Stowe, et al. (2022) | UK | Mixed (BNT162b2/mRNA-1273/ChAdOx1) | EC -> H | 18-64 | 14-174 | 0.593 [ 0.539, 0.641] |
| Buchan, et al. (2022a) | Canada | Mixed (≥1 mRNA Vaccine) | H/D | ≥18 | 7-59 | 0.550 [-1.060, 0.900] |
| Adams, et al. (2022) | USA | Mixed (BNT162b2/mRNA-1273/Janssen) | H | ≥18 | 14-150 | 0.530 [ 0.330, 0.670] |
| Pooled VE in adults: (Q = 23.82, df = 6, p < .01; $I^2$ = 75.5%, $\tau^2$ = 1.16e-01) | | | | | | 0.707 [ 0.592, 0.789] |
| Pooled VE in adults across all vaccine types: (Q = 28.24, df = 9, p < .01; $I^2$ = 67.1%, $\tau^2$ = 5.84e-02) | | | | | | 0.699 [ 0.628, 0.756] |
| Pooled VE in all age groups across all vaccine types: (Q = 123.06, df = 15, p < .01; $I^2$ = 87.0%, $\tau^2$ = 1.63e-01) | | | | | | 0.669 [ 0.583, 0.738] |
| Test for VE Difference in All across Vaccine Types: $Q_M$ = 0.80, df = 1, p = 0.37 | | | | | | |

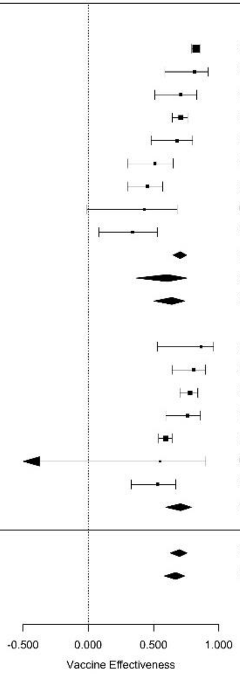

sFigure 5 Short-term vaccine effectiveness of full dose against Severe Events

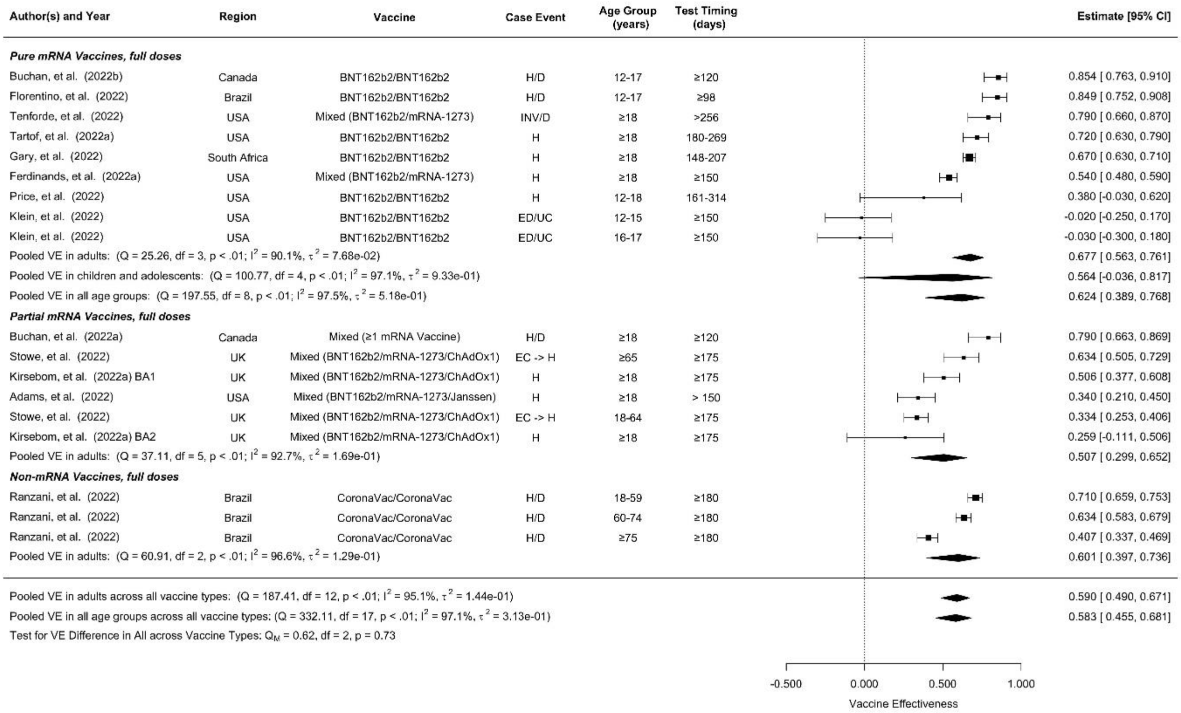

sFigure 6 Long-term vaccine effectiveness of full dose against Severe Events

| Author(s) and Year | Region | Vaccine | Case Event | Age Group (years old) | Test Timing (days) | Estimate [95% CI] |
|---|---|---|---|---|---|---|
| ***Pure mRNA Vaccines, booster dose, short term*** | | | | | | |
| Ferdinands, et al. (2022a) | USA | Mixed (BNT162b2/mRNA-1273) | H | ≥18 | <60 | 0.910 [0.880, 0.930] |
| Natarajan, et al. (2022) | USA | Mixed (BNT162b2/mRNA-1273) | H | ≥18 | 7-120 | 0.900 [0.880, 0.910] |
| Tartof, et al. (2022a) | USA | BNT162b2/BNT162b2/BNT162b2 | H | ≥18 | <90 | 0.850 [0.800, 0.890] |
| Grewal, et al. (2022a) | Canada | BNT162b2/BNT162b2/mRNA-1273 | H/D | ≥60 | 16-98 | 0.810 [0.670, 0.890] |
| Grewal, et al. (2022a) | Canada | Mixed (BNT162b2/mRNA-1273) | H/D | ≥60 | <84 | 0.800 [0.720, 0.860] |
| Grewal, et al. (2022a) | Canada | BNT162b2/BNT162b2/BNT162b2 | H/D | ≥60 | 24-112 | 0.770 [0.670, 0.830] |
| Pooled VE in adults: (Q = 38.43, df = 5, p < .01; $I^2$ = 86.7%, $\tau^2$ = 1.33e-01) | | | | | | 0.853 [0.798, 0.893] |
| ***Partial mRNA Vaccines, booster dose, short term*** | | | | | | |
| Stowe, et al. (2022) | UK | Mixed (BNT162b2/mRNA-1273/ChAdOx1) | EC -> H | ≥65 | 7-69 | 0.935 [0.919, 0.947] |
| Ranzani, et al. (2022) | Brazil | CoronaVac/CoronaVac/BNT162b2 | H/D | 18-59 | 8-59 | 0.921 [0.902, 0.937] |
| Kirsebom, et al. (2022a) BA1 | UK | Mixed (BNT162b2/mRNA-1273/ChAdOx1) | H | ≥18 | 7-69 | 0.892 [0.866, 0.913] |
| Ranzani, et al. (2022) | Brazil | CoronaVac/CoronaVac/BNT162b2 | H/D | 60-74 | 8-59 | 0.884 [0.862, 0.902] |
| Kirsebom, et al. (2022a) BA2 | UK | Mixed (BNT162b2/mRNA-1273/ChAdOx1) | H | ≥18 | 7-69 | 0.872 [0.786, 0.923] |
| Stowe, et al. (2022) | UK | Mixed (BNT162b2/mRNA-1273/ChAdOx1) | EC -> H | 18-64 | 7-69 | 0.823 [0.807, 0.837] |
| Ranzani, et al. (2022) | Brazil | CoronaVac/CoronaVac/BNT162b2 | H/D | ≥75 | 8-59 | 0.773 [0.731, 0.808] |
| Pooled VE in adults: (Q = 142.46, df = 6, p < .01; $I^2$ = 95.7%, $\tau^2$ = 1.84e-01) | | | | | | 0.881 [0.834, 0.914] |
| ***Non-mRNA Vaccines, booster dose, short term*** | | | | | | |
| Ranzani, et al. (2022) | Brazil | CoronaVac/CoronaVac/CoronaVac | H/D | 18-59 | 8-59 | 0.860 [0.717, 0.931] |
| Ranzani, et al. (2022) | Brazil | CoronaVac/CoronaVac/CoronaVac | H/D | 60-74 | 8-59 | 0.807 [0.686, 0.881] |
| Natarajan, et al. (2022) | USA | Janssen/Janssen | H | ≥18 | 7-120 | 0.670 [0.520, 0.770] |
| Ranzani, et al. (2022) | Brazil | CoronaVac/CoronaVac/CoronaVac | H/D | ≥75 | 8-59 | 0.477 [0.124, 0.688] |
| Pooled VE in adults: (Q = 12.29, df = 3, p < .01; $I^2$ = 78.6%, $\tau^2$ = 2.35e-01) | | | | | | 0.730 [0.537, 0.843] |
| ***Pure mRNA Vaccines, booster dose, long term*** | | | | | | |
| Tenforde, et al. (2022) | USA | Mixed (BNT162b2/mRNA-1273) | INV/D | ≥18 | >60 | 0.940 [0.880, 0.970] |
| Grewal, et al. (2022a) | Canada | mRNA-1273/mRNA-1273/mRNA-1273 | H/D | ≥60 | 70-150 | 0.810 [0.740, 0.860] |
| Ferdinands, et al. (2022a) | USA | Mixed (BNT162b2/mRNA-1273) | H | ≥18 | 120-149 | 0.780 [0.670, 0.850] |
| Grewal, et al. (2022a) | Canada | Mixed (BNT162b2/mRNA-1273) | H/D | ≥60 | ≥84 | 0.770 [0.690, 0.820] |
| Tartof, et al. (2022a) | USA | BNT162b2/BNT162b2/BNT162b2 | H | ≥18 | ≥90 | 0.550 [0.280, 0.710] |
| Pooled VE in adults: (Q = 23.98, df = 4, p < .01; $I^2$ = 91.1%, $\tau^2$ = 3.85e-01) | | | | | | 0.801 [0.646, 0.888] |
| ***Partial mRNA Vaccines, booster dose, long term*** | | | | | | |
| Ranzani, et al. (2022) | Brazil | CoronaVac/CoronaVac/BNT162b2 | H/D | 60-74 | ≥60 | 0.904 [0.891, 0.915] |
| Ranzani, et al. (2022) | Brazil | CoronaVac/CoronaVac/BNT162b2 | H/D | 18-59 | ≥60 | 0.902 [0.883, 0.917] |
| Stowe, et al. (2022) | UK | Mixed (BNT162b2/mRNA-1273/ChAdOx1) | EC -> H | ≥65 | ≥105 | 0.862 [0.812, 0.898] |
| Kirsebom, et al. (2022a) BA1 | UK | Mixed (BNT162b2/mRNA-1273/ChAdOx1) | H | ≥18 | ≥105 | 0.804 [0.756, 0.843] |
| Ranzani, et al. (2022) | Brazil | CoronaVac/CoronaVac/BNT162b2 | H/D | ≥75 | ≥60 | 0.785 [0.761, 0.806] |
| Stowe, et al. (2022) | UK | Mixed (BNT162b2/mRNA-1273/ChAdOx1) | EC -> H | 18-64 | ≥105 | 0.605 [0.512, 0.681] |
| Kirsebom, et al. (2022a) BA2 | UK | Mixed (BNT162b2/mRNA-1273/ChAdOx1) | H | ≥18 | ≥105 | 0.565 [0.384, 0.693] |
| Adams, et al. (2022) | USA | Mixed (BNT162b2/mRNA-1273/Janssen) | H | ≥18 | > 120 | 0.390 [0.220, 0.530] |
| Pooled VE in adults: (Q = 327.30, df = 7, p < .01; $I^2$ = 98.2%, $\tau^2$ = 4.74e-01) | | | | | | 0.780 [0.643, 0.864] |
| ***Non-mRNA Vaccines, booster dose, long term*** | | | | | | |
| Ranzani, et al. (2022) | Brazil | CoronaVac/CoronaVac/CoronaVac | H/D | 18-59 | ≥60 | 0.820 [0.609, 0.917] |
| Ranzani, et al. (2022) | Brazil | CoronaVac/CoronaVac/CoronaVac | H/D | 60-74 | ≥60 | 0.761 [0.708, 0.804] |
| Ranzani, et al. (2022) | Brazil | CoronaVac/CoronaVac/CoronaVac | H/D | ≥75 | ≥60 | 0.514 [0.446, 0.573] |
| Pooled VE in adults: (Q = 37.97, df = 2, p < .01; $I^2$ = 94.2%, $\tau^2$ = 2.22e-01) | | | | | | 0.705 [0.473, 0.835] |
| Pooled VE in adults across all vaccine types, Short Term: (Q = 235.81, df = 16, p < .01; $I^2$ = 94.9%, $\tau^2$ = 2.45e-01) | | | | | | 0.848 [0.804, 0.881] |
| Pooled VE in adults across all vaccine types, Long Term: (Q = 539.10, df = 15, p < .01; $I^2$ = 97.0%, $\tau^2$ = 3.72e-01) | | | | | | 0.776 [0.694, 0.836] |

Test for Short Term VE Difference in Adults across Vaccine Types: $Q_M$ = 7.82, df = 2, p = 0.02
Test for Long Term VE Difference in Adults across Vaccine Types: $Q_M$ = 0.57, df = 2, p = 0.75

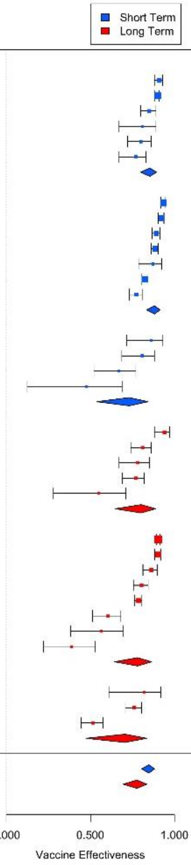

sFigure 7 Short-term and long-term vaccine effectiveness of first booster dose against Severe Events

**Acronyms in the Supplementary Figures:**

SI: symptomatic infection

AI: all infection

D: death

H: hospitalization

ED/UC: emergency department (ED) or urgent care (UC) encounter

ED: emergency department admission

EC→H: hospital admissions from emergency care

H/D: hospitalization or death

ICU: intensive care unit (ICU) admission

NCH: noncritical hospitalization

INV/D: invasive mechanical ventilation/death

INV: invasive ventilation

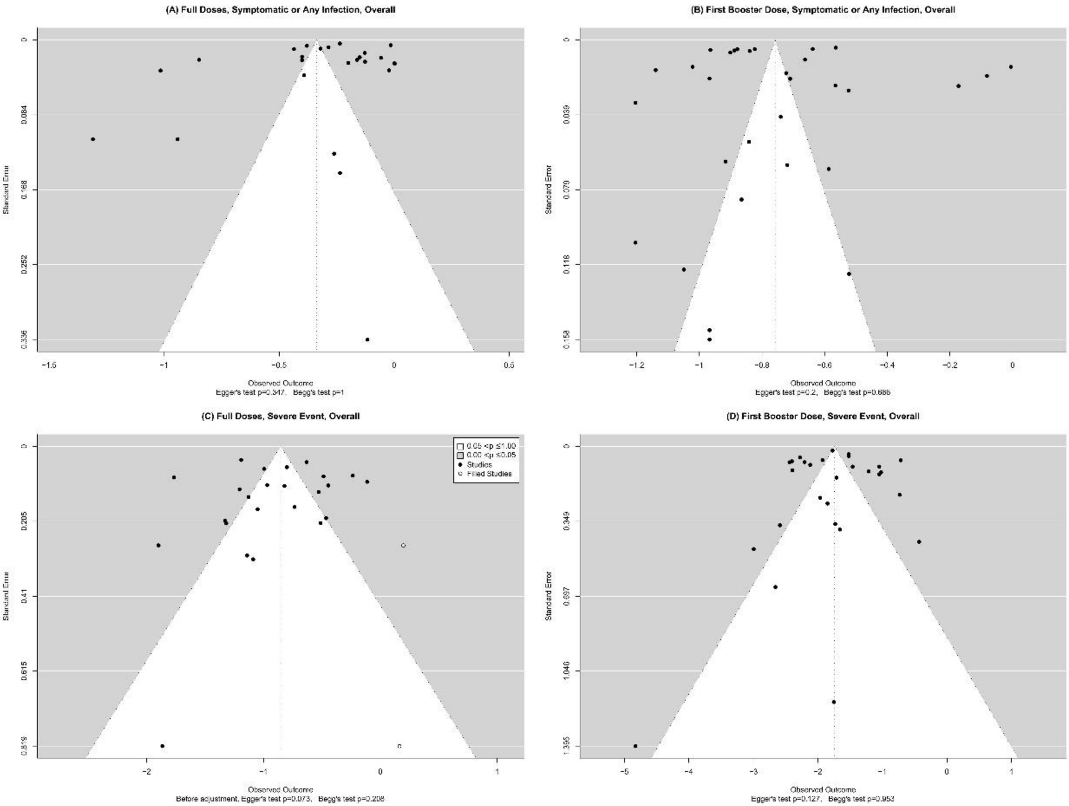

sFigure 8 Funnel plots for meta-analyses of overall VE estimates: (A) full doses against symptomatic infection or any infection; (B) first booster doses against symptomatic infection or any infection; (C) full doses against severe events; (D) first booster doses against severe events.

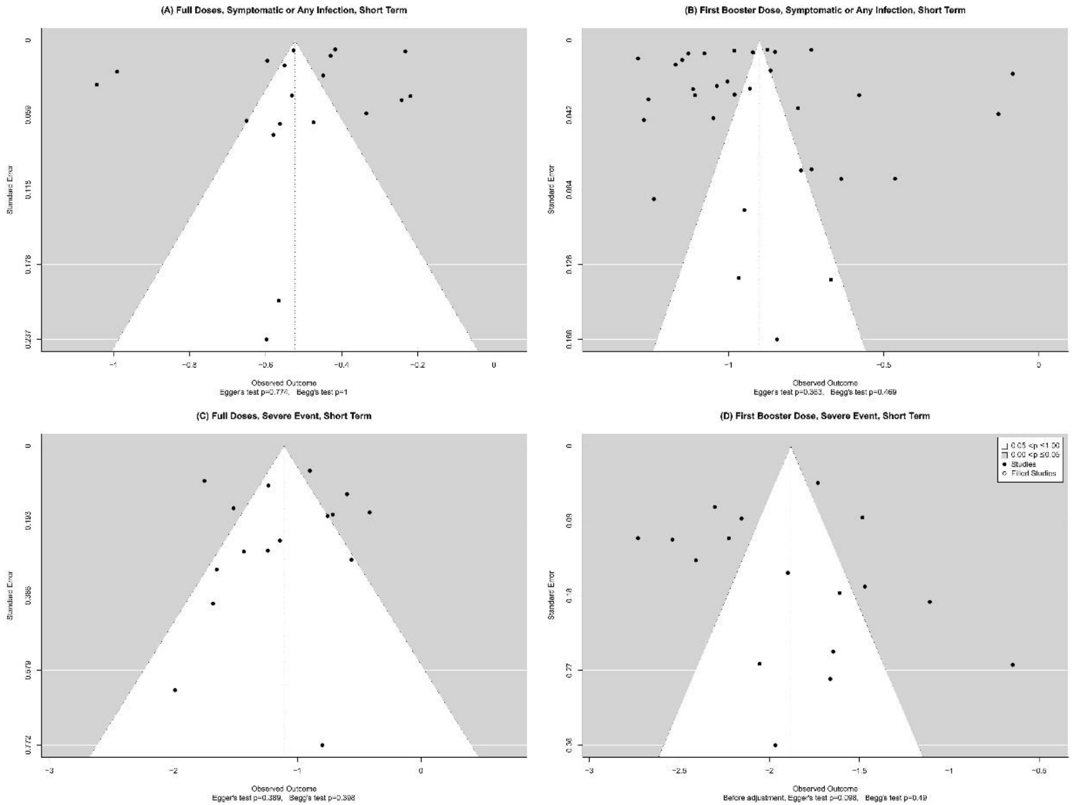

sFigure 9 Funnel Plots for meta-analyses of short-term VE estimates: (A) full doses against symptomatic infection or any infection; (B) first booster doses against symptomatic infection or any infection; (C) full doses against severe events; (D) first booster doses against severe events

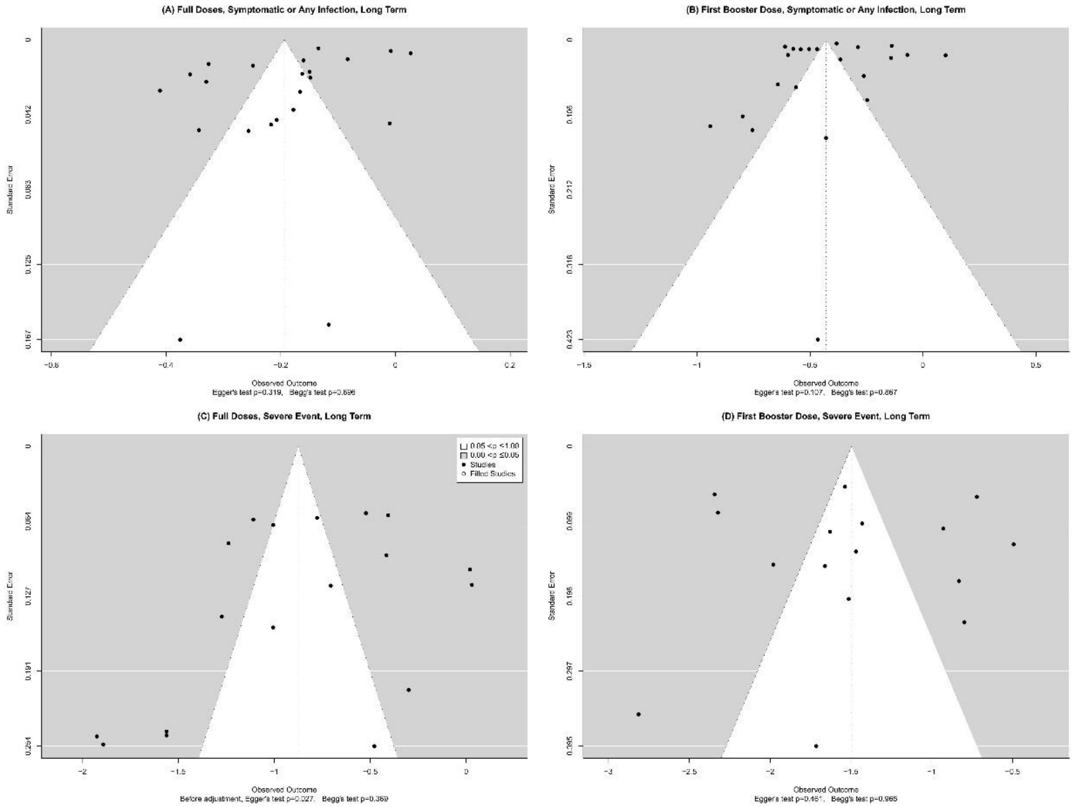

sFigure 10 Funnel Plots for meta-analyses of long-term VE estimates: (A) full doses against symptomatic infection or any infection; (B) first booster doses against symptomatic infection or any infection; (C) full doses against severe events; (D) first booster doses against severe events

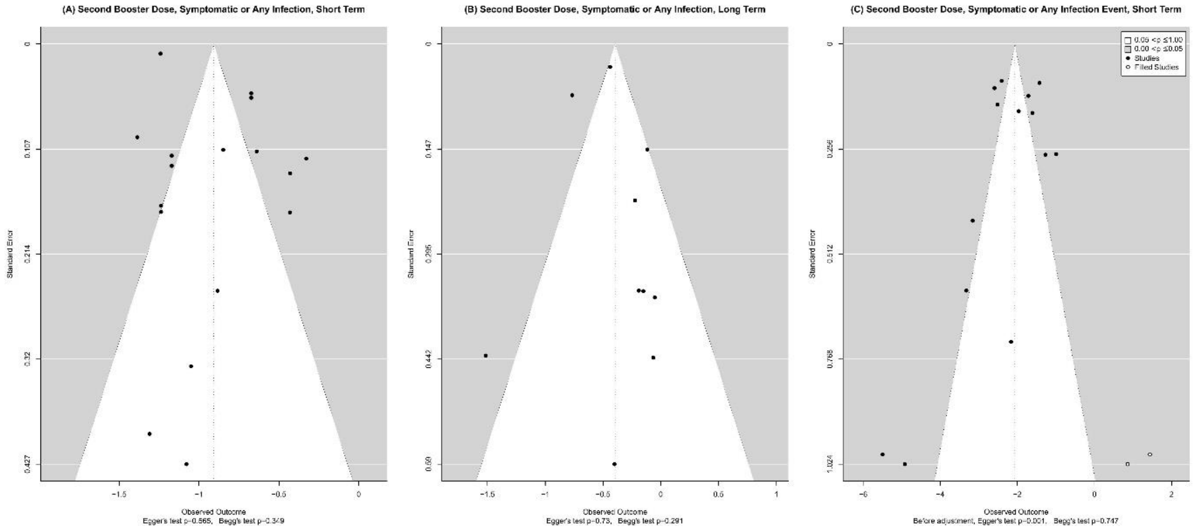

sFigure 11 Funnel Plots for meta-analyses of VE estimates of the second booster dose against: (A) symptomatic infection or any infection in the short term; (B) symptomatic infection or any infection in the long term; (C) severe events in the short term. For the second booster, VE estimates were only available for adults.

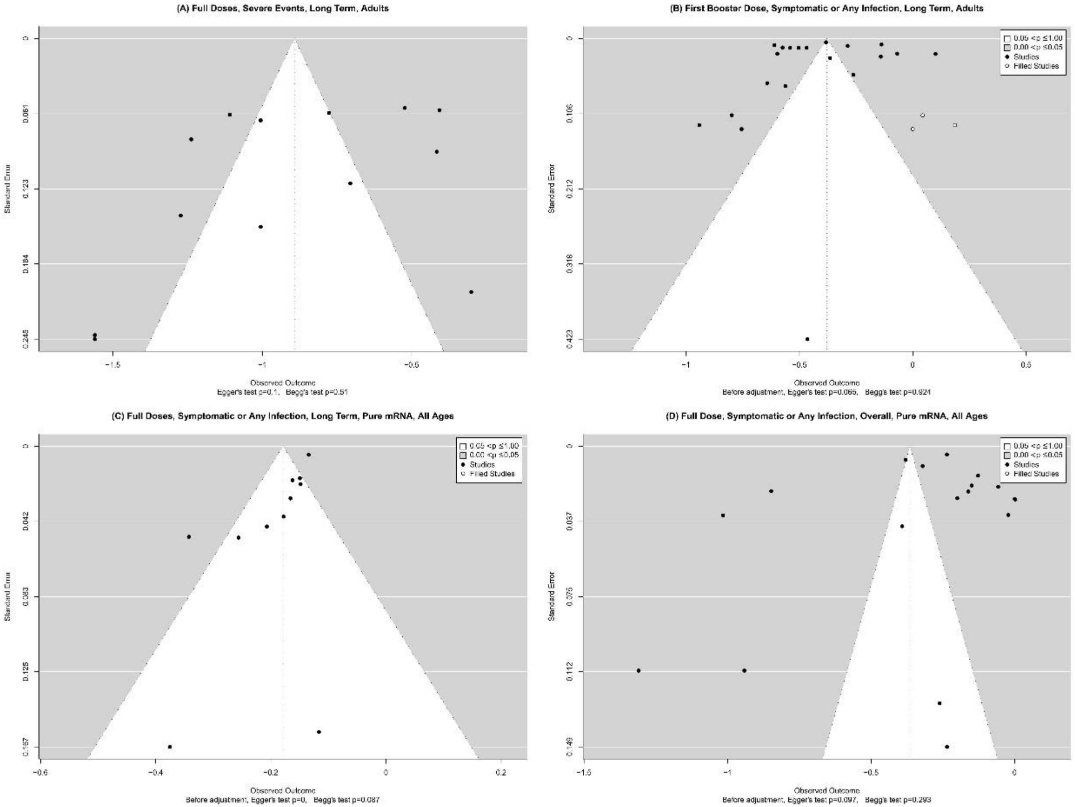

sFigure 12 Funnel plots for meta-analyses of VE estimates in subgroups defined by age group and vaccine type that show publication bias (any p-value < 0.1)

#### Searching Strategies

**Full Doses and Booster Search Strategy:**

Search conducted time: June 27th, 2022

Publication time: November 26th, 2021, to June 27th, 2022, if a date can be specified; 2021 to 2022 if the year is the most specific scope.

Key words:

#1 (SARS-CoV-2) OR (COVID-19) OR (2019nCoV)
#2 (vaccine) OR (vaccination)
#3 (effectiveness) OR (efficacy)
#4 (test-negative) OR (case-control) OR (cohort study)
#5 Omicron
#6 (infected) OR (infection) OR (hospitalization) OR (hospital admission)

Note for #4, although our main interest is test-negative case-control design studies, we also searched for the other two possible study types to reduce the number of false negative results.

| Database | Number of results |
|---|---|
| PubMed | 82 |
| Web of Science | 23 |
| Embase | 89 |
| Scopus | 721 |
| #1 AND #2 AND #3 AND #4 AND #5 AND #6 searched in all fields | |

Cochrane library only supports up to 5 search terms

| Database | Number of results |
|---|---|
| Cochrane Library | 3 |
| #1 AND #2 AND #3 AND #4 AND #5 searched in all fields | |

Preprint databases used different searching rules and limitations

| Database | Number of results |
|---|---|
| medRxiv | 115 |
| bioRxiv | 6 |
| for term "SARS-CoV-2 COVID-19 2019nCoV vaccine effectiveness test-negative case-control cohort study Omicron infection hospitalization" and posted between "November 26th, 2021 and June 27th, 2022" | |

Google Scholar is a supplementary search source and can give many false positive results, so only the first ten pages of most relevant results will be screened.

| Google Scholar | Top 100 out of 4,290 |
|---|---|
| ((SARS-CoV-2) OR (COVID-19) OR (2019nCoV)) AND ((vaccine) OR (vaccination)) AND ((effectiveness) OR (efficacy)) AND ((test-negative) OR (case-control) OR (cohort study)) AND (Omicron) AND ((infected) OR (infection) OR (hospitalization) | |

**Second Booster Search Strategy:**
Search conducted time: Jan 08th, 2023
Publication time: November 26th, 2021, to Jan 08th, 2023, if a date can be specified; 2021 to 2023 if the year is the most specific scope.

Key words:
#1 (SARS-CoV-2) OR (COVID-19) OR (2019nCoV)
#2 (second booster) OR (additional booster) OR (fourth dose)
#3 (effectiveness) OR (efficacy)
#4 (test-negative) OR (case-control) OR (cohort study)
#5 Omicron
#6 (infected) OR (infection) OR (hospitalization) OR (hospital admission)

Note for #4, although our main interest is test-negative case-control design studies, we also searched for the other two possible study types to reduce the number of false negative results.

| Database | Number of results |
|---|---|
| PubMed | 56 |
| Web of Science | 22 |
| Embase | 55 |
| Scopus | 1015 |
| #1 AND #2 AND #3 AND #4 AND #5 AND #6 searched in all fields | |

Cochrane library only supports up to 5 search terms

| Database | Number of results |
|---|---|
| Cochrane Library | 9 |
| #1 AND #2 AND #3 AND #4 AND #5 searched in all fields | |

Preprint databases used different searching rules and limitations

| Database | Number of results |
|---|---|
| medRxiv | 149 |
| bioRxiv | 7 |
| for term "SARS-CoV-2 COVID-19 2019nCoV second booster effectiveness test-negative case-control study Omicron infection hospitalization" and posted between "November 26th, 2021 and Sep 05th, 2022" | |

Google Scholar is a supplementary search source and can give many false positive results, so only the first ten pages of most relevant results will be screened.

| Google Scholar | Top 100 out of 4,110 |
|---|---|
| ((SARS-CoV-2) OR (COVID-19) OR (2019nCoV)) AND ((second booster) OR (additional booster) OR (fourth dose)) AND ((effectiveness) OR (efficacy)) AND ((test-negative) OR (case-control) OR (cohort study)) AND (Omicron) AND ((infected) OR (infection) OR (hospitalization) | |

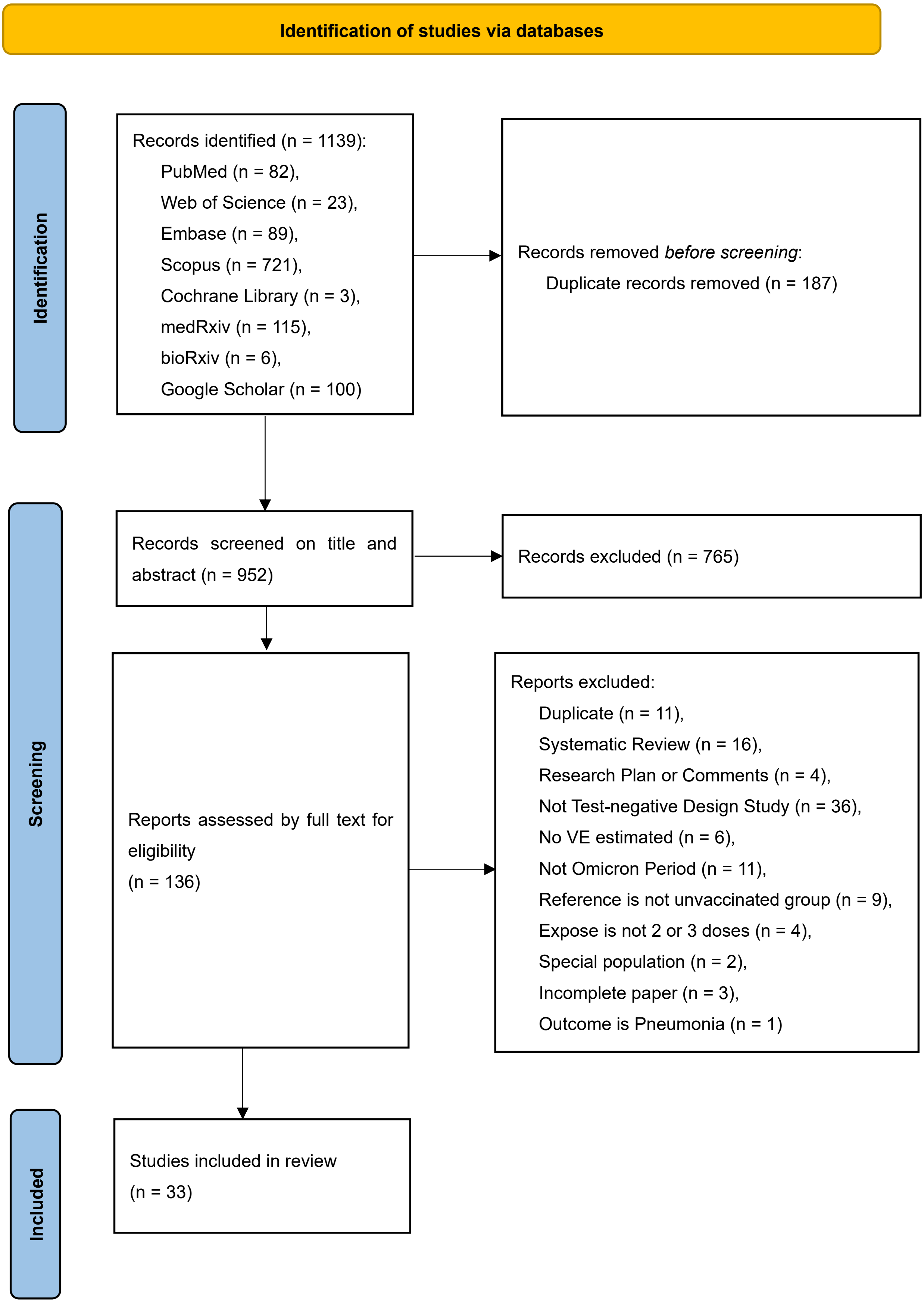

sFigure 13 PRISMA flow diagram for full doses and booster

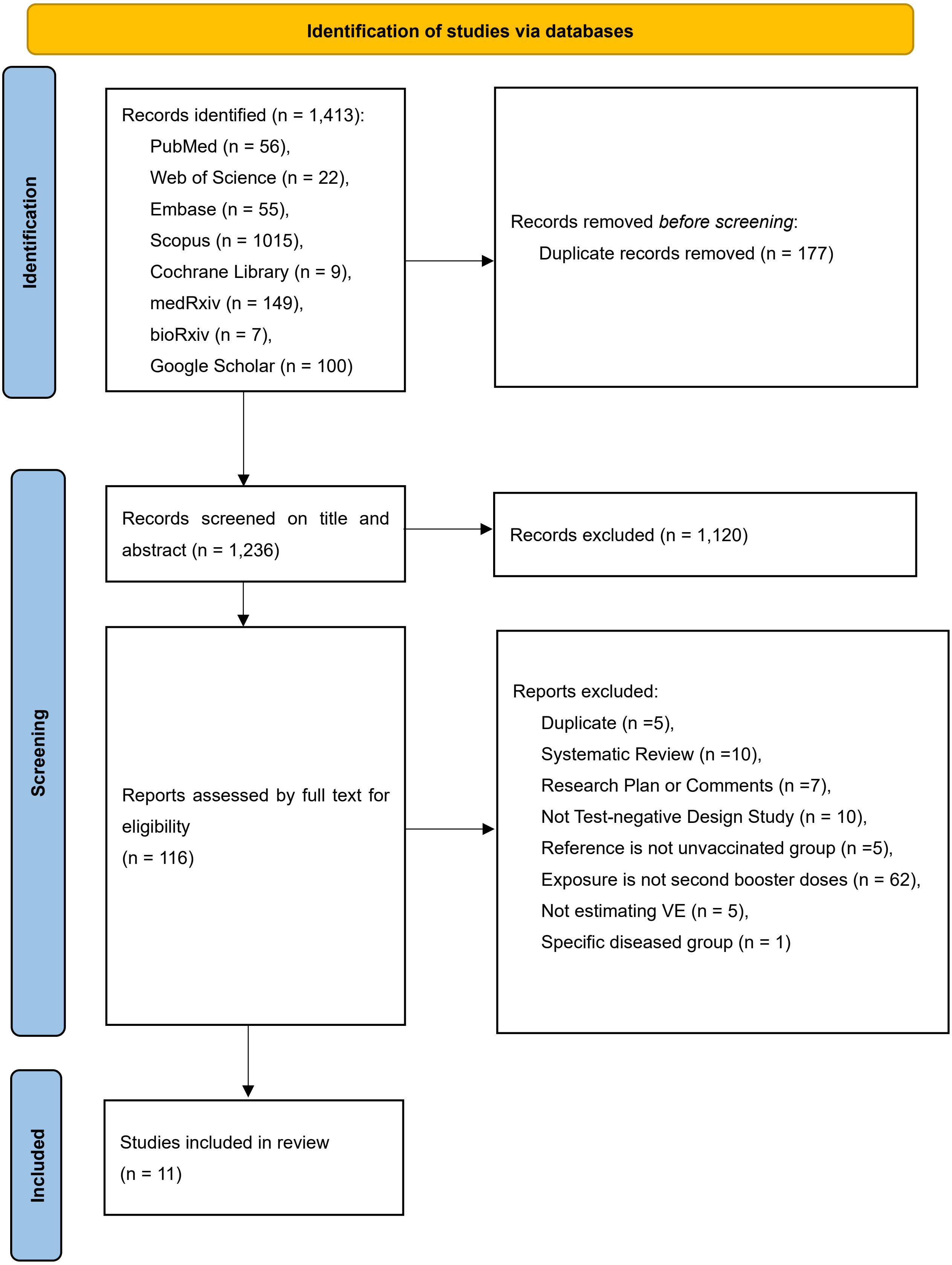

sFigure 14 PRISMA flow diagram for second booster
